# Widespread genetic effect heterogeneity impacts bias and power in nonlinear Mendelian randomization

**DOI:** 10.64898/2026.04.17.26351133

**Authors:** Jiongming Wang, Jean Morrison

## Abstract

Mendelian randomization (MR) uses genetic variants as instrumental variables to infer causal relationships between complex traits. Standard MR can be used to estimate an average causal effect at the population level, and typically assumes a linear exposure-outcome relationship. Recently, several methods for estimating nonlinear effects have been developed. However, many have been found to produce spurious empirical findings when subjected to negative control analyses. We propose that this poor performance may be attributable to heterogeneity in variant-exposure associations. We demonstrate that heterogeneous genetic effects on exposure lead to biased estimates, poor coverage, and inflated type I error in control function and stratification-based methods. In contrast, two-stage least squares (TSLS) methods are robust to such heterogeneity, but suffer from low precision and low power in some circumstances. We show that a statistical test for heterogeneity can be used to guide the choice of nonlinear MR methods. Using UK Biobank data, we reassess the causal effects of BMI, vitamin D, and alcohol consumption on blood pressure, lipid, C-reactive protein, and age (negative control). We find strong evidence of heterogeneity for all three exposures, and also recapitulate previous results that control function and stratification-based methods are prone to false positives. Finally, using nonparametric TSLS, we identify evidence of nonlinear causal effects of BMI on HDL cholesterol, triglycerides, and C-reactive protein; however, specific estimates of the shape of these relationships are imprecise. Altogether, our results suggest that common nonlinear MR methods are unreliable in the presence of realistic levels of heterogeneity, and that more methodological development is required before practically useful nonlinear MR is feasible.

## 2 Introduction

Identifying causal relationships among complex traits has long been a fundamental question in public health and biomedical research. Randomized control trails are the gold standard for identifying and quantifying causal effects, but they are often infeasible due to ethical challenges and cost limitations. Alternatively, Mendelian randomization (MR), as an instrumental variable approach, can estimate causal effects from observational associations between modifiable exposure and health-related outcome traits[1]. Mendelian randomization can provide valid causal estimates in the presence of unmeasured confounding, enabling its wide applications in epidemiological and human genetic research. It has provided important insights into disease etiologies and has informed causal relationships across a wide range of traits[2–8].

Standard MR methods implicitly assume that the causal effect of a quantitative exposure on the outcome is linear on an additive or multiplicative scale. However, nonlinear associations are commonly observed, suggesting a potential for nonlinear causal effects[9–14]. For example, nonlinear J- or U-shaped associations have been documented and reproduced between habitual alcohol consumption and the risk of cardiovascular diseases[9, 13]. However, it remains controversial whether these observational associations reflect underlying nonlinear causal effects, or are due to confounding factors, reverse causality, or selection bias. Several MR methods for estimating nonlinear causal relationships have been proposed, including stratification-based methods[15–17], control function methods[18, 19], and nonlinear extensions of two-stage least squares (TSLS)[20–25]. All nonlinear Mendelian randomization methods require individual-level data.

Nonlinear MR methods have been broadly implemented in biobank analyses[12, 13]. However, recent studies have indicated that the stratification-based nonlinear MR methods can produce spurious empirical findings[26–28], leading to retractions and corrections of previous analyses using these methods[29– 31]. One potential source of these errors is heterogeneity of instrument-exposure associations within the population[28, 32, 33], although the underlying mechanisms remain poorly understood, particularly for non-stratification-based approaches. In this article, we show that instrument-exposure association heterogeneity results in violation of identifying conditions for stratification-based and control function methods. This violation induces biased and miscalibrated causal estimates as well as inflated rates of false identification of causal effects and of false rejection of causal linearity. We further show both theoretically and through simulation that heterogeneity does not invalidate TSLS methods; however, these methods can be underpowered compared to stratification-based and control function methods in the absence of heterogeneity. We investigate the use of diagnostic tests for heterogeneity, and show that pre-estimation screening can reduce the risk of false positives. Finally, we reassess the causal effects of BMI, vitamin D, and alcohol consumption on blood pressure, lipids, and inflammation traits in UK Biobank. We find empirical evidence of instrument-exposure effect heterogeneity for these exposures and compare their estimates using different nonlinear MR methods.

## 3 Methods

### 3.1 Nonlinear MR and instrument-exposure effect heterogeneity

Nonlinear MR methods assume that exposure, *X*, outcome, *Y*, and instrumental variables (genetic variants), *Z* are related by the structural equations

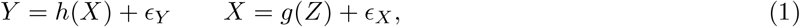

where *h*(·) and *g*(·) are unknown, potentially nonlinear functions, and *ϵ*_*Y*_ and *ϵ*_*X*_ are residual errors with mean zero. In nonlinear MR analysis, we seek to estimate the causal effect of increasing *X* on *Y*, which is given by the derivative 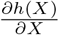.

We review three classes of nonlinear MR estimators, two-stage least squares (TSLS), control function, and stratification-based methods. All three approaches rely on two core instrumental variable assumptions, relevance and exogeneity. Relevance requires that the instrument, *Z*, is correlated with the exposure, *X*. Exogeneity requires that the instrument is uncorrelated with the error term *ϵ*_*Y*_ . In MR literature, the exogeneity assumption is usually divided into two assumptions, that the instrument is not affected by unmeasured confounders (exchangeability), and that the instrument affects the outcome only through the exposure (exclusion restriction).

TSLS methods approximate the causal function, *h*, and instrument-exposure association, *g*, using linear combinations of basis functions, *η*(·) and *ζ*(·), respectively[18, 34],

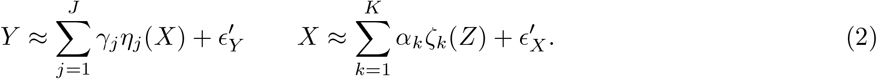

In the first stage, we fit *J* regressions, one for each basis function in the outcome model,

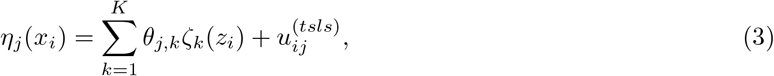

obtaining coefficient estimates 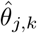 for *j* ∈ 1, …, *J, k* ∈ 1, …, *K*. From these estimates, we compute predicted values, 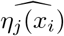. In the second stage, we fit the outcome regression,

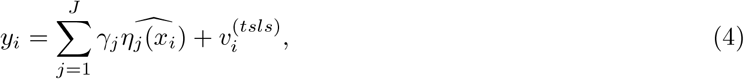

to obtain estimates, 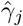, of the causal parameters. In practice, *η*_*j*_(·) and *ζ*_*k*_(·) are often specified as polynomial or spline basis functions. Conventional TSLS methods can be extended by adopting more flexible modeling techniques, including nonparametric methods[24, 25] and deep learning approaches[20, 23]. Validity of TSLS methods requires only the core relevance and exogeneity assumptions. In the context of (2), relevance requires that *E*(*I*(*X*)*I*(*Z*)^*T*^) is of full rank, where *I*(*X*) = (*η*_1_(*X*), …, *η*_*J*_ (*X*))^*T*^ and *I*(*Z*) = (*ζ*_1_(*Z*), …, *ζ*_*K*_(*Z*))^*T*^, and exogeneity requires that 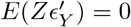. TSLS methods do not require homogeneity in the instrument-exposure relationship. They only require that each of the basis functions in the outcome model can be independently instrumented, meaning that they can be predicted from *Z* and that these predictions are not collinear.

Like TSLS, control function approaches also estimate the causal effect of *X* on *Y* using a two-stage procedure[18]. Using the same approximation as in (2), we first fit the exposure regression

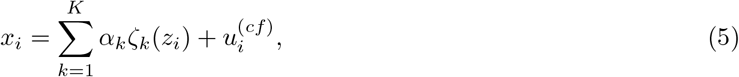

to obtain estimates, 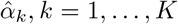, fitted values, 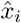, and residuals, 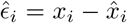. In the second stage, we regress the outcome on the exposure as well as the residuals from the first stage,

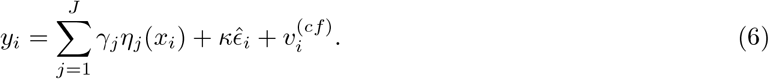

The resulting coefficient estimates, 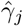, *j* ∈ 1, …, *J*, correspond to the causal parameters in (2). The PolyMR method[19] extends this approach by including higher-order residual terms in second-stage regression to accommodate more complex confounding structures. For correct identification, control function methods require the two core assumptions and, additionally, that the instrument is uncorrelated with the residual error of the instrument-exposure association, i.e., 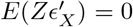, and that the conditional expectation 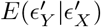 is a linear function of 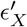 and 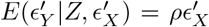, for some value of *ρ*. In other words, the control function approach is only valid if the model for the relationship between the instrument and exposure fully captures all dependence of the exposure on the instrument. By contrast, in the TSLS methods, the instrument-exposure relationship does not need to be correctly specified. Guo and Small show that the control function estimator can be viewed as a TSLS estimator with an augmented set of instrumental variables, with the additional instruments being valid only when the instrument-exposure model is correctly specified.

Instrument-exposure effect heterogeneity results in violation of the additional assumptions required by control function approaches, when the source of heterogeneity is unknown. For example, suppose that *X* is related to a single instrument *Z* through the structural equation

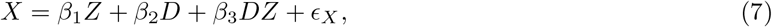

where *D* is an unobservable source of heterogeneity such as overall health or unmeasured environmental factors. If *D* is unobservable, it is impossible to correctly specify the instrument-exposure model. If we fit a simple linear regression of *X* on *Z*, the resulting residuals will not be independent of *Z*.

In stratification-based methods, the sample is first partitioned and then standard MR is performed within each stratum. Stratifying on the value of exposure itself would introduce bias due to colliding. Therefore, stratification-based methods first regress the exposure on instrument, as in the first stage of the control function approach to obtain residuals. Individuals are then stratified based on these residuals, and separate causal effect is computed within each stratum. These estimators can be viewed as approximating the underlying causal function with a piecewise function. The residual stratification method[15] creates strata as quantiles of the exposure residuals. Like the control function approach, residual stratification requires that the residual used for stratification is independent of the exposure, and additionally that relevance and exogeneity hold within each stratum. Doubly-ranked stratification methods[16, 17] relax this assumption and require that the ranks of *X*_*i*_(*z*) are invariant with respect to *z* (i.e., rank-preserving assumption).

Instrument-exposure effect heterogeneity leads to violations of assumptions for all stratification-based methods. Like the control function approach, the original residual stratification method fails whenever the instrument-exposure relationship is misspecified, which we have shown occurs in the presence of heterogeneity. Heterogeneity also results in violations of the rank-preserving assumption. For example, suppose that the exposure and instrument are related by (7), and that both *Z* and *D* are binary. Then for *Z* = 0, values of *X*_*i*_(*Z* = 0) are ordered according to *β*_2_*D*_*i*_ + *ϵ*_*X,i*_, while values of *X*_*i*_(*Z* = 1) are ordered according to *β*_1_ + (*β*_2_ + *β*_3_)*D*_*i*_ + *ϵ*_*X,i*_, which are not necessarily the same for some values of *β*_1_, *β*_2_, and *β*_3_.

Stratification-based methods assume a single instrument, such as a polygenic score (PGS), while TSLS and control function methods can be applied using either a single summary instrument or multiple instruments[18]. However, using multiple instruments may be computationally expensive or, when *J* is large, can result in an excessive number of parameters.

### 3.2 Simulation study

#### 3.2.1 Generating individual-level genotype, exposure and outcome data

To evaluate the impact of instrument-exposure effect heterogeneity on nonlinear MR estimators, we simulated individual-level data from a range of scenarios. We first simulated genotypes for 1, 000 independent causal variants. For each individual, *i*, and variant, *j*, we sampled *G*_*i,j*_ from a Binomial(2, *f*_*j*_) distribution, with the allele frequency, *f*_*j*_, drawn from a Beta(1, 5) distribution. We converted the genotypes to the standardized scale by centering them around 2*f*_*j*_ and dividing by 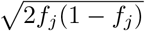. We additionally simulated one continuous confounder of the exposure and outcome, *U*_*i*_, from a standard normal distribution.

We simulated exposure values for each individual as

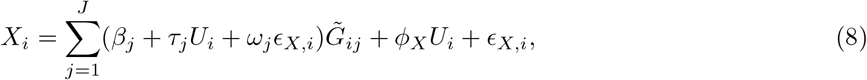

where 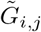 is the standardized genotype, *ϕ*_*X*_ is the confounder effect on the exposure, *ϵ*_*X,i*_ is a normally distributed environmental component with variance chosen so that the total variance of *X*_*i*_ is one, and *β*_*j*_, *τ*_*j*_, and *ω*_*j*_ are effect size parameters that vary between scenarios.

We simulated the outcome for each individual as

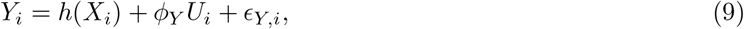

where *h* encodes the potentially nonlinear relationship between the exposure and outcome, *ϕ*_*Y*_ is the confounder effect on the outcome, and *ϵ*_*Y,i*_ is a normally distributed environmental component of the outcome with variance chosen so that the total variance of *Y*_*i*_ is one.

For each simulation scenario, we specify the total heritability of *X* as 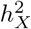. We also specify the heritability of *X* attributable to the instrument-confounder interaction, 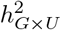, and the heritability of *X* attributable to the instrument-environment interaction, 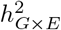, and define 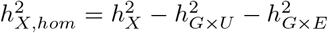. Homogeneous effect sizes, *β*_*j*_, for each variant were drawn from a Normal 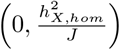 distribution, following the genomewide complex trait analysis (GCTA) framework[35, 36].

The heterogeneous components of the variant association may be correlated with the homogeneous effect, *β*_*j*_, due to pathway-driven effects, or may be independent of the homogeneous effect[37]. To capture these mechanisms, we drew *τ*_*j*_ and *ω*_*j*_ from two-component mixture distributions,

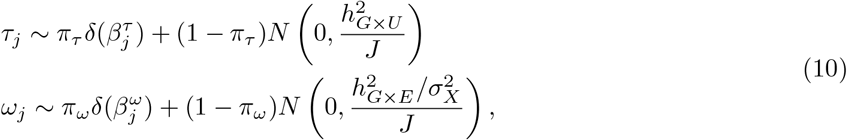

where *δ*(*x*_0_) indicates the density with a point mass at *x*_0_, 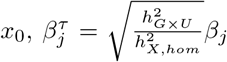 and 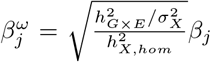, and 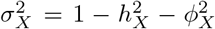 is the variance of *ϵ*_*X,i*_. We refer to the two components of these distributions as the directional component and balanced component.

We considered simulation scenarios with no heterogeneous effects 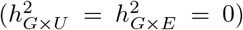, variant-by-confounder heterogeneity only (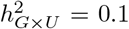 and 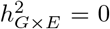), or variant-by-environment heterogeneity only (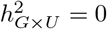 and 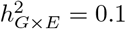). Across all scenarios, we fixed the homogeneous heritability at 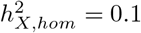. In settings with heterogeneity, we considered *π*_*τ*_, *π*_*ω*_ ∈ {0, 0.33, 0.66, 1}, representing balanced, mixed (balanced- or directional-driven), and directional heterogeneity structures, respectively.

We considered four specifications of *h*(·) in equation (9), no effect (*h*(*X*) = 0), linear (*h*(*X*) = 0.05*X*), symmetric quadratic (*h*(*X*) = 0.05*X*^2^), and asymmetric quadratic (*h*(*X*) = 0.05*X*^2^ +0.05*X*). In this simulation, the exposure is symmetrically distributed with mode at zero, so symmetry of the causal effect function around zero corresponds to symmetry of the causal effect function around the mode of the exposure. The confounder-exposure and confounder-outcome effects were set as 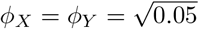 across all simulations.

In each replicate, we simulated two independent samples of equal size, one for polygenic score calculation (discovery cohort), and the other for nonlinear MR estimation (evaluation cohort). We considered sample sizes of 50, 000 and 100, 000 to represent low-power and high-power settings, respectively.

#### 3.2.2 Implementing nonlinear MR methods

We applied all nonlinear MR methods using a single summary instrument, a PGS computed using summary statistics from the discovery cohort. To compute the PGS, we first estimated variant-exposure associations using linear regression and selected variants with *p <* 5 × 10^−8^ in the discovery cohort. We then computed the score in the evaluation cohort as 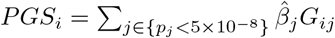.

Methods compared included TSLS methods Deep IV[20], Quantile IV[23], and NPIV[24, 25]; control function method PolyMR[19]; and stratification-based methods residual stratification[15], and doubly-ranked stratification[16] (see Note S1). We additionally included parametric versions of the TSLS and control function approaches, using polynomial bases with *J* = 2 (quadratic) and *J* = 3 (cubic) in both stages as well as versions of these approaches in which the polynomial degree was selected using AIC, considering degrees 1 to 10. Parametric TSLS was implemented using the `AER` package, while the parametric control function approach was implemented with base R functions. Finally, we fitted the linear TSLS estimator as a benchmark.

In each simulation, we implemented the Breusch-Pagan test[38, 39] to screen for evidence of instrument-exposure effect heterogeneity, using the `lmtest` R package with default options.

#### 3.2.3 Evaluation metrics

We evaluated nonlinear MR methods in terms of bias, mean squared error (MSE), and coverage for estimates of the causal effect, 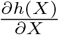, as well as type I error, and power to reject the global null of no causal effect at any value of *X* and to reject the hypothesis that the effect is linear. Bias, MSE, and coverage were averaged over the observed values of the exposure in the evaluation cohort. NPIV does not directly provide tests of the global and linearity null hypotheses. However, Chen et al. suggest rejecting the global null at level *α* if at least one 1 − *α* confidence interval excludes 0 over the observed values of the exposure, and rejecting the linearity null if no single value is contained in all 1 − *α* confidence intervals. For PolyMR, we tested for overall and nonlinear effects using likelihood ratio tests implemented in the `PolyMR` package. For the residual stratification and doubly-ranked stratification methods, two nonlinearity tests are available from the `SUMnlmr` package[40], the quadratic test, which compares a linear and quadratic exposure-outcome model, and the Cochran’s Q test, which is nonparametric. For both of these methods, we computed an overall effect test by combining stratum-specific p-values using the Cauchy combination association test[41, 42]. For polynomial TSLS and polynomial control function approaches, we tested the overall effect using a Wald test of the null hypothesis that all coefficients were equal to zero. We tested for nonlinearity by testing that coefficients of nonlinear exposure terms were equal to zero. DeepIV and QuantileIV do not provide standard uncertainty quantification or hypothesis testing. Although Legault et al. suggest bagging-based confidence intervals, these are not directly comparable to conventional inferential procedures. For these methods, we only evaluated bias and MSE.

### 3.3 Analysis in UK Biobank

UK Biobank is a prospective cohort study containing genetic, lifestyle and health information collected from about 500, 000 participants between 40 and 69 years old across the United Kingdom recruited in 2006-2010[43]. We restricted our analysis to 332, 305 unrelated individuals who self-reported as white British.

We evaluated the causal effects of BMI, vitamin D, and alcohol consumption on systolic and diastolic blood pressure, low density lipoprotein (LDL) cholesterol, high density lipoprotein (HDL) cholesterol, triglycerides, total cholesterol, and C-reactive protein, as well as effects of all three exposures on age, which is treated as a negative control. BMI was computed from height and weight measured at the initial visit (unit: kg/m^2^). Vitamin D level was measured by chemiluminescent immunoassay (CLIA) analysis (unit: nmol/L) and was log transformed. We calculated alcohol consumption by weighting the self-reported average weekly intake of different types of alcohol and converting it into American standard measure (unit: drink/week)[13], added one, and then log transformed the resulting values. All exposures were then standardized to have mean zero and variance one.

Systolic and diastolic blood pressure were measured using an automated device reading (unit: mmHg). LDL cholesterol, HDL cholesterol, triglycerides, and total cholesterol were measured using enzymatic protective selection, enzyme immunoinhibition, GPO-POD, and CHO-POD assays, respectively (unit: mmol/L). C-reactive protein was measured using high-sensitivity immunoturbidimetric analysis (unit: mg/L). All outcomes were measured at the initial visit. We applied a log transformation to triglycerides and C-reactive protein, and standardized all outcome traits to have mean zero and variance one.

We used 73 instruments previously identified in nonlinear MR analyses for BMI (Burgess et al.), 21 instruments for vitamin D (Sofianopoulou et al.), and 93 instruments for alcohol consumption (Kassaw et al.). Instrument-exposure association estimates were obtained from previous GWAS. For BMI, we downloaded summary statistics from the OpenGWAS database[45] (OpenGWAS ID: ieu-a-835), based on the original study performed by the Genetic Investigation of ANthropometric Traits (GIANT) consortium[46], which did not include UK Biobank participants. For vitamin D, we used summary statistics generated from the previous nonlinear MR analyses[30], which was performed in UK Biobank. For alcohol consumption, we used summary statistics from [47], which meta-analyzed over 940, 000 individuals, including approximately 300, 000 UK Biobank participants. We then extracted genotypes for each variant in the UK Biobank cohort, and computed the polygenic scores for each individual using the external association estimates as weights. We further standardized the scores to have mean zero and variance one, and used these standardized scores as instruments in downstream analysis.

#### 3.3.1 Statistical analysis

We implemented six nonlinear MR methods across all 24 exposure-outcome pairs, NPIV, quadratic TSLS, PolyMR, quadratic control function, residual stratification, and doubly-ranked stratification. DeepIV and QuantileIV were not implemented, as they do not provide estimates of uncertainty or hypothesis tests. We applied NPIV using both tensor-product and generalized B-spline polynomial bases, and implemented PolyMR both with and without model selection. We also assessed instrument-exposure effect heterogeneity using the Breusch-Pagan test. All analyses except for age were adjusted for age, age squared, sex, age-by-sex, age-squared-by-sex, birthplace, recruitment center, genotyping array, and the top 40 genetic principal components. All of these covariates except those involving age were included in the age analysis. NPIV and PolyMR cannot directly accommodate covariates. For these methods, we regressed the exposure and outcome on the covariates and used the residuals from these models in place of the original exposure and outcome values. For all other nonlinear MR methods, covariates were included in both regression stages. We also included covariates in the Breusch-Pagan test. All analyses were conducted using complete-case data.

## 4 Results

### 4.1 Comparison of nonlinear MR methods in simulations

Under homogeneity or fully balanced heterogeneity, including both *G* × *U* and *G* × *E* settings, all nonlinear MR methods except DeepIV produce unbiased estimates, and achieve average coverage at or above the nominal level (Figures 1, S1, and S2). The bias of deep learning methods is potentially due to the use of cross validation for model selection[25]. Additionally, in these scenarios, all methods except PolyMR with model selection control the type I error for both the overall and nonlinearity tests (Figures 3A and 4A). The inflated type I error of PolyMR with model selection is likely attributable to failing to account for model selection in the subsequent significance tests.

**Figure 1.**
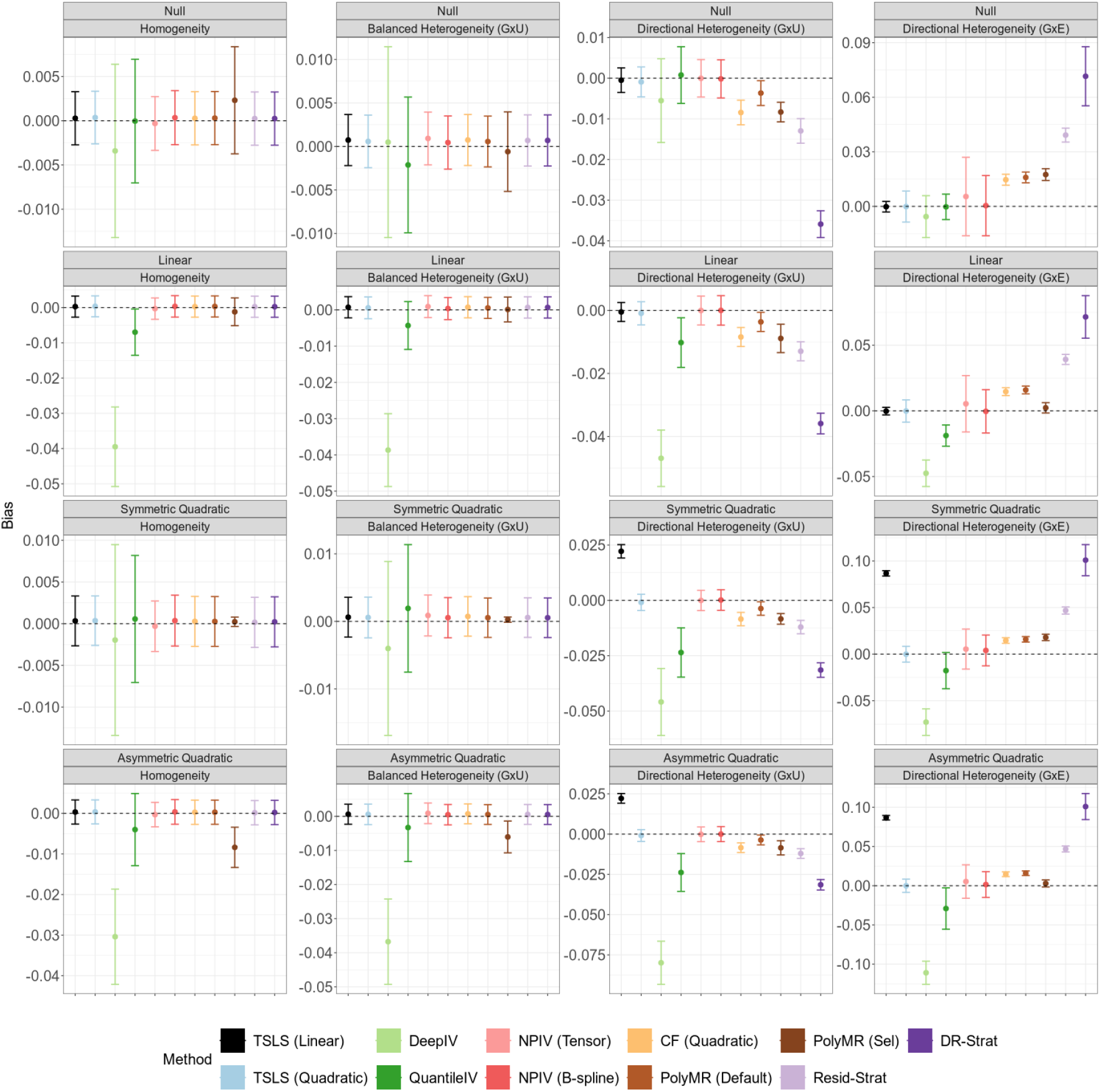
Average bias of nonlinear MR methods under instrument-exposure effect homogeneity or heterogeneity. Points indicate the average bias across 100 replicates, and error bars indicate 95% confidence intervals for the mean. Simulation sample size is 100, 000. In the balanced heterogeneity (*G* × *U*) setting, *π*_*τ*_ = 0. In the directional heterogeneity (*G* × *U*) and (*G* × *E*) settings, *π*_*τ*_ = 1 or *π*_*ω*_ = 1, respectively. TSLS, two-stage least squares; NPIV, NPIV with tensor-product basis (Tensor) or generalized B-spline polynomial basis (B-spline); CF, control function; PolyMR (Default), PolyMR without model selection; PolyMR (Sel), PolyMR with model selection; Resid-Strat, residual stratification; DR-Strat, doubly-ranked stratification.

Consistent with theoretical predictions, control function and stratification-based methods are biased, have reduced coverage, and inflated type I error rates for both tests under directional heterogeneity. This inflation is more pronounced under *G* × *U* heterogeneity than under *G* × *E* heterogeneity, and increases as exposure GWAS power increases (Figures S4 and S5). One exception to this pattern is that doubly-ranked stratification with the quadratic test does control the type I error for the nonlinearity test under *G* × *E* directional heterogeneity, but not under *G* × *U* directional heterogeneity (Figure 4A).

The quadratic TSLS and NPIV methods remain unbiased in the presence of directional heterogeneity, maintain proper coverage, and control the type I error. However, these methods can have low power and low precision in many settings. In particular, when there is no or only balanced heterogeneity and the causal effect is symmetric quadratic, the TSLS-based methods have almost no power to identify an overall causal effect or the evidence of nonlinearity (Figures 3B and 4B). This reflects a failure of the relevance assumption, as the predictions 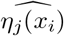 tend to be collinear in these scenarios. NPIV shows the lowest power across all methods and scenarios, reflecting the imprecision of its estimates, while quadratic TSLS has similar power to linear TSLS to identify an overall causal effect. Both quadratic TSLS and NPIV also have low power to identify nonlinear causal effects, with power improving in the presence of heterogeneity. Additionally, for both tests, power increases as the exposure GWAS becomes more powerful (Figures S4 and S5).

Although the quadratic control function approach is biased and has poorly calibrated tests under directional heterogeneity, it has the lowest MSE across all methods and scenarios (Figure 2). Quadratic TSLS performs best among the methods that remain unbiased in the presence of heterogeneity, and it outperforms linear TSLS when there is directional heterogeneity and the causal shape is nonlinear. All other methods have higher MSE than linear TSLS in the presence of directional heterogeneity across all causal effect shapes.

**Figure 2.**
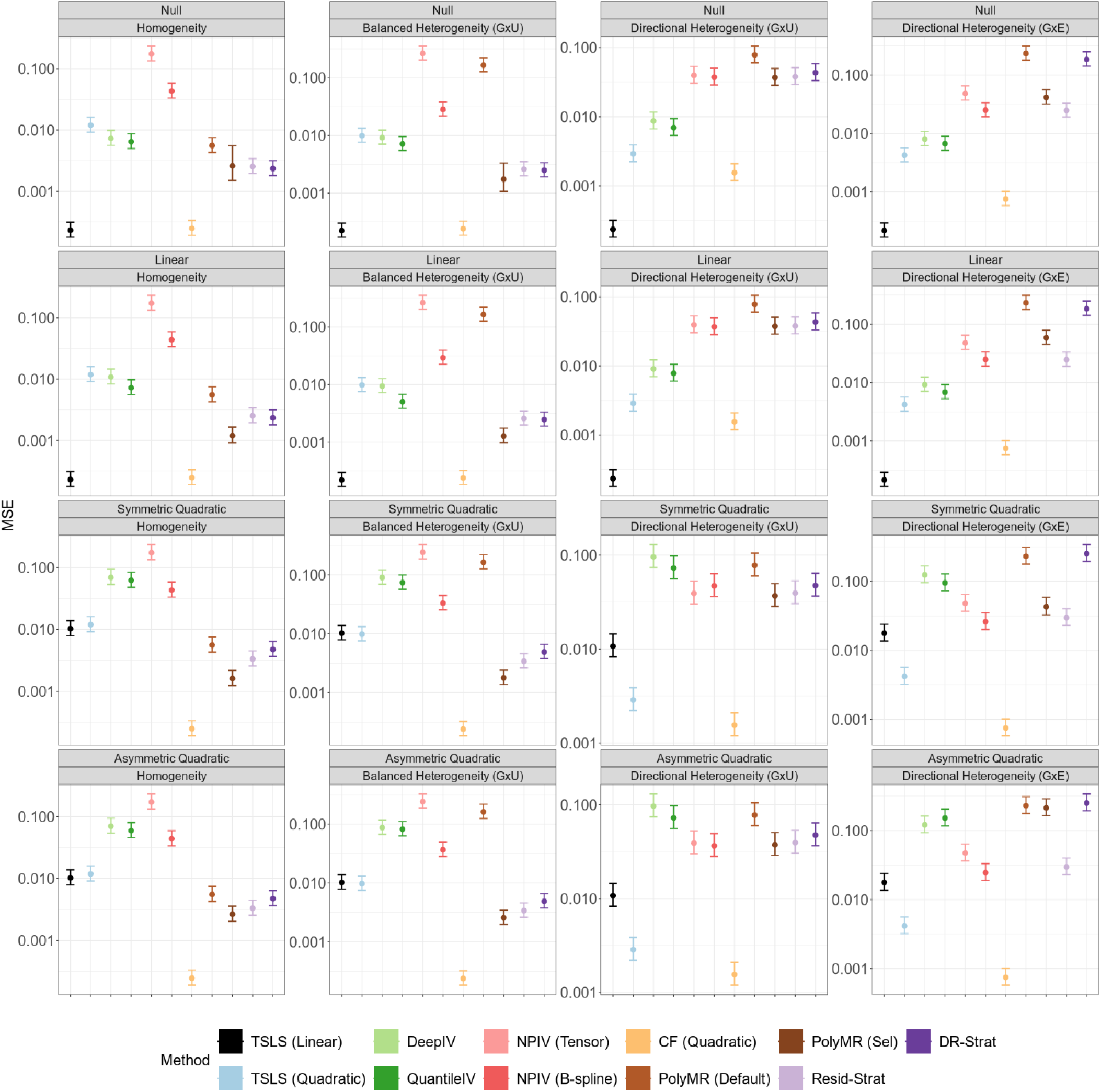
Average mean squared error (MSE) of nonlinear MR methods under instrument-exposure effect homogeneity or heterogeneity. Points indicate the average MSE across 100 replicates, and error bars indicate 95% confidence intervals for the mean. Simulation sample size is 100, 000. In the balanced heterogeneity (*G* × *U*) setting, *π*_*τ*_ = 0. In the directional heterogeneity (*G* × *U*) and (*G* × *E*) settings, *π*_*τ*_ = 1 or *π*_*ω*_ = 1, respectively. TSLS, two-stage least squares; NPIV, NPIV with tensor-product basis (Tensor) or generalized B-spline polynomial basis (B-spline); CF, control function; PolyMR (Default), PolyMR without model selection; PolyMR (Sel), PolyMR with model selection; Resid-Strat, residual stratification; DR-Strat, doubly-ranked stratification.

**Figure 3.**
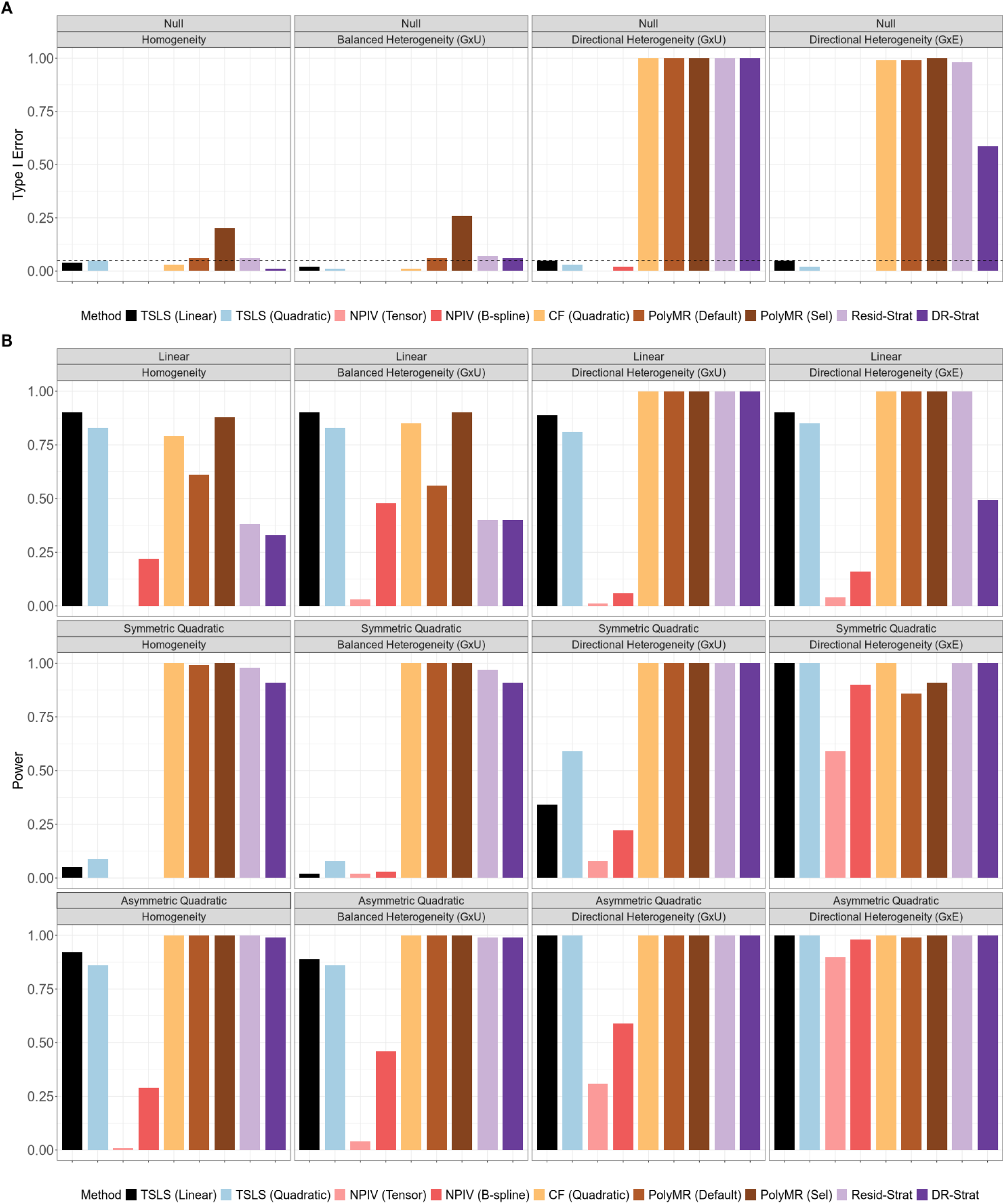
Type I error and power for overall-effect testing of nonlinear MR methods. (A) Type I error. (B) Power. Type I error and power were calculated from *N* = 100, 000 simulated individuals per replicate, and summarized across 100 replicates. In the balanced heterogeneity (*G* × *U*) setting, *π*_*τ*_ = 0. In the directional heterogeneity (*G* × *U*) and (*G* × *E*) settings, *π*_*τ*_ = 1 or *π*_*ω*_ = 1, respectively. TSLS, two-stage least squares; NPIV, NPIV with tensor-product basis (Tensor) or generalized B-spline polynomial basis (B-spline); CF, control function; PolyMR (Default), PolyMR without model selection; PolyMR (Sel), PolyMR with model selection; Resid-Strat, residual stratification; DR-Strat, doubly-ranked stratification.

**Figure 4.**
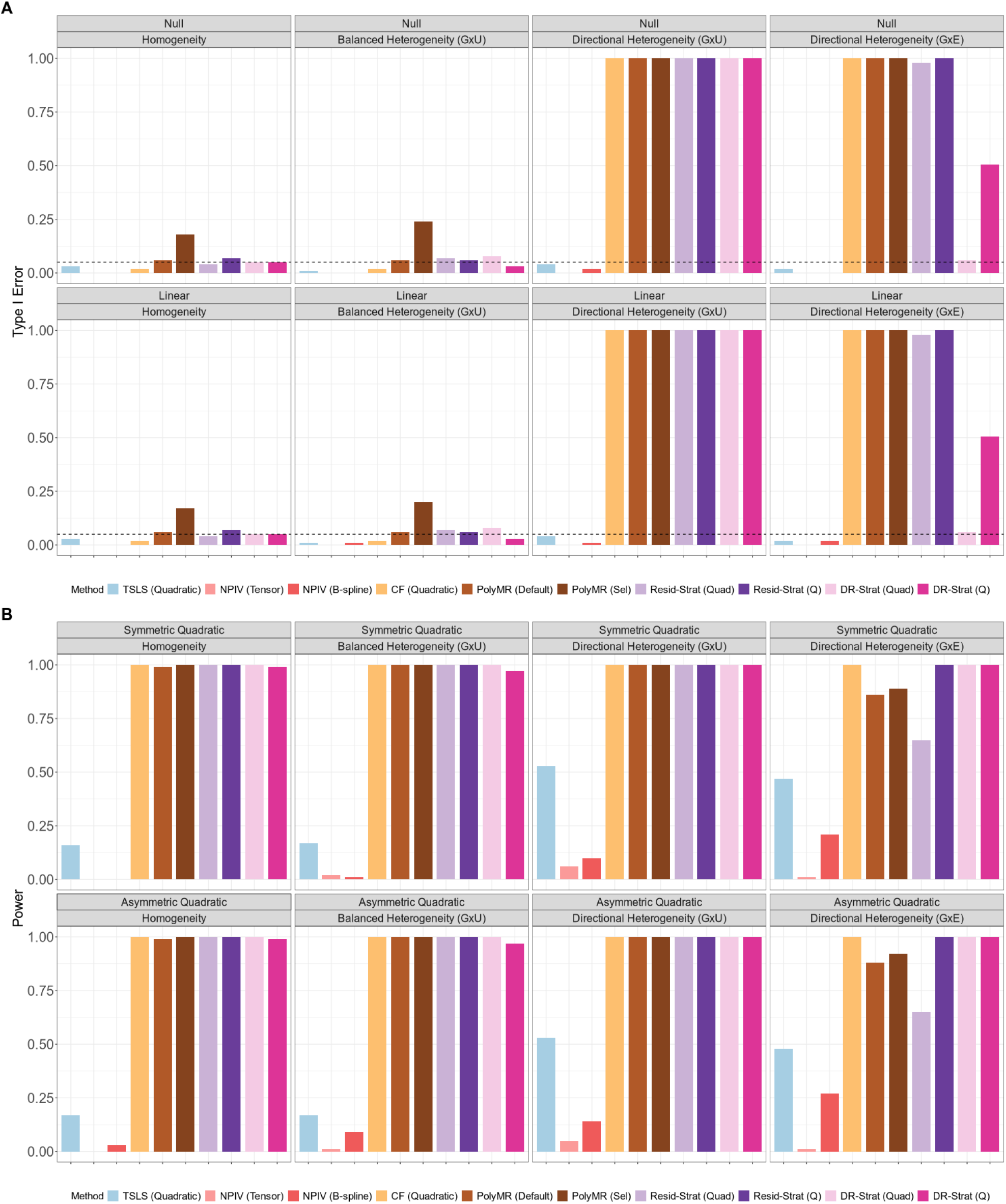
Type I error and power for nonlinearity testing of nonlinear MR methods. (A) Type I error. (B) Power. Type I error and power were calculated from *N* = 100, 000 simulated individuals per replicate, and summarized across 100 replicates. In the balanced heterogeneity (*G* × *U*) setting, *π*_*τ*_ = 0. In the directional heterogeneity (*G* × *U*) and (*G* × *E*) settings, *π*_*τ*_ = 1 or *π*_*ω*_ = 1, respectively. TSLS, two-stage least squares; NPIV, NPIV with tensor-product basis (Tensor) or generalized B-spline polynomial basis (B-spline); CF, control function; PolyMR (Default), PolyMR without model selection; PolyMR (Sel), PolyMR with model selection; Resid-Strat, residual stratification; DR-Strat, doubly-ranked stratification; Quad, quadratic test; Q, Cochran’s Q test.

Under directional heterogeneity, TSLS with an insufficient polynomial degree can be biased, and exhibit reduced coverage because it fails to capture the underlying higher-order structure (Figures S1 and S2), while including unnecessary higher-degree terms can reduce power (Figures S4 and S5). More generally, polynomial degree misspecifications, whether too low or too high, can increase MSE for TSLS under directional heterogeneity (Figure S3). In contrast, control function is less sensitive to polynomial degree specification. We also observed a bias-variance tradeoff due to model selection for polynomial TSLS and PolyMR. Model selection reduced MSE for these methods in some settings, but introduced additional bias and decreased coverage (Figures S1-S3). Model selection also inflated type I error rates for PolyMR and polynomial control function, even when these methods achieved nominal type I error control without model selection (Figures S4 and S5).

We stratified the overall test p-values for nonlinear MR methods in null scenarios with no causal effect by the p-value of the Breusch-Pagan test for heterogeneity. Figure 5A presents quantile-quantile plots with p-values pooled across heterogeneity settings. We found that all methods controlled the type I error amongst simulations with non-significant Breusch-Pagan test statistics (*p >* 0.1). This suggests that the Breusch-Pagan test could serve as an effective screening tool before applying nonlinear MR methods. We observed the same pattern for tests of nonlinearity (Figure 5B).

**Figure 5.**
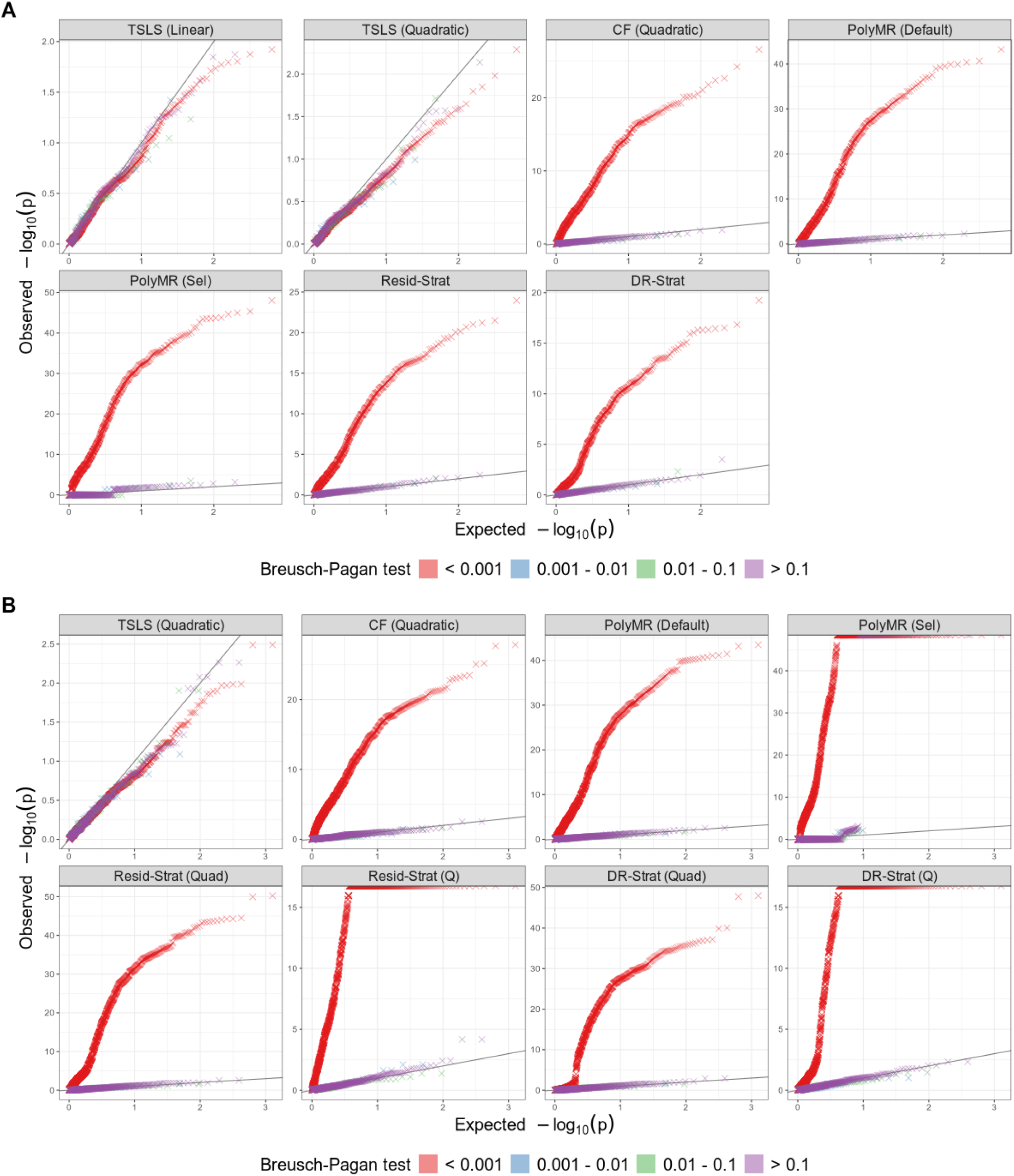
Quantile–Quantile plots for overall-effect and nonlinearity testing of nonlinear MR methods. (A) Overall-effect testing, evaluated under the null hypothesis of no causal effect. (B) Nonlinearity testing, evaluated under the null hypothesis of no causal effect or a linear effect. P-values were calculated from *N* = 100, 000 simulated individuals, and pooled across all homogeneity and heterogeneity scenarios over 100 replicates. TSLS, two-stage least squares; CF, control function; PolyMR (Default), PolyMR without model selection; PolyMR (Sel), PolyMR with model selection; Resid-Strat, residual stratification; DR-Strat, doubly-ranked stratification; Quad, quadratic test; Q, Cochran’s Q test.

### 4.2 Assessing causal effects of BMI, vitamin D, and alcohol consumption in UK Biobank

In the UK Biobank, Breusch-Pagan tests provide strong evidence of instrument-exposure effect heterogeneity across all three exposures (*p <* 1 × 10^−15^ for all exposures). This suggests that control function and stratification-based methods will be biased and susceptible to false discoveries in this setting.

Across all 24 exposure-outcome trait pairs, linear MR (linear TSLS) identifies 15 nominally significant causal effects (Table 1; Figure S7). None of the nonlinear TSLS methods identifies an overall effect that is not detected by linear TSLS. This pattern is consistent with simulation results, which show that linear TSLS is more powerful than nonlinear TSLS for identification of an overall causal effect. Among the nonlinear TSLS methods, quadratic TSLS identifies the largest number of overall effects and differs from linear TSLS only for the effects of alcohol consumption on C-reactive protein and age. None of the TSLS methods identifies an overall causal effect for the negative control outcome age, though notably, the linear TSLS method does identify a nominally significant effect of alcohol consumption on age (*p* = 0.04). This suggests there may be some additional sources of bias, such as uncorrected population stratification, affecting these estimates.

**Table 1:** Linear and nonlinear Mendelian randomization results of overall-effect estimation and inference for BMI, vitamin D, and alcohol consumption on blood pressure, lipid, C-reactive protein, and age. ★ indicates significant overall-effect test results at *α* = 0.05, and —indicates non-significant results. Arrows indicate the direction (sign) of the estimated causal effect from the linear MR analysis.

| Exposure | Outcome | Linear MR | Nonlinear MR |  |  |  |  |  |  |  |
| --- | --- | --- | --- | --- | --- | --- | --- | --- | --- | --- |
|  |  | TSLS-L | TSLS-Q | NPIV-T | NPIV-B | CF-Q | PolyMR-D | PolyMR-S | Strat-RS | Strat-DR |
| BMI | SBP | ★↑ | ★ | ★ | ★ | ★ | ★ | ★ | ★ | ★ |
|  | DBP | ★↑ | ★ | ★ | ★ | ★ | ★ | ★ | ★ | ★ |
|  | LDL | ★↓ | ★ | ★ | ★ | ★ | ★ | ★ | ★ | ★ |
|  | HDL | ★↓ | ★ | ★ | ★ | ★ | ★ | ★ | ★ | ★ |
|  | TG | ★↑ | ★ | ★ | ★ | ★ | ★ | ★ | ★ | ★ |
|  | TC | ★↓ | ★ | ★ | ★ | ★ | ★ | ★ | ★ | ★ |
|  | CRP | ★↑ | ★ | ★ | ★ | ★ | ★ | ★ | ★ | ★ |
|  | Age | —↓ | — | — | — | ★ | ★ | ★ | ★ | ★ |
| Vitamin D | SBP | —↓ | — | — | — | ★ | — | ★ | — | — |
|  | DBP | —↓ | — | — | — | ★ | — | — | — | — |
|  | LDL | —↓ | — | — | — | ★ | ★ | — | ★ | — |
|  | HDL | ★↑ | ★ | — | — | ★ | ★ | ★ | ★ | ★ |
|  | TG | ★↓ | ★ | — | — | ★ | ★ | ★ | ★ | ★ |
|  | TC | —↑ | — | — | — | ★ | ★ | — | ★ | — |
|  | CRP | —↓ | — | — | — | — | — | ★ | — | — |
|  | Age | —↑ | — | — | — | ★ | — | — | — | — |
| Alcohol | SBP | ★↑ | ★ | ★ | ★ | ★ | ★ | ★ | ★ | ★ |
|  | DBP | ★↑ | ★ | ★ | ★ | ★ | ★ | ★ | ★ | ★ |
|  | LDL | —↓ | — | — | — | ★ | — | — | ★ | ★ |
|  | HDL | ★↑ | ★ | — | — | ★ | ★ | ★ | ★ | ★ |
|  | TG | ★↓ | ★ | ★ | ★ | ★ | ★ | ★ | ★ | ★ |
|  | TC | —↑ | — | — | — | ★ | — | ★ | — | ★ |
|  | CRP | ★↑ | — | — | — | ★ | ★ | ★ | ★ | ★ |
|  | Age | ★↓ | — | — | — | ★ | — | ★ | ★ | — |

Abbreviations: TSLS-L, linear two-stage least squares; TSLS-Q, quadratic two-stage least squares; NPIV-T, NPIV with tensor-product basis; NPIV-B, NPIV with generalized B-spline polynomial basis; CF-Q, quadratic control function; PolyMR-D, PolyMR without model selection (default); PolyMR-S, PolyMR with model selection; Strat-RS, residual stratification; Strat-DR, doubly-ranked stratification; SBP, systolic blood pressure; DBP, diastolic blood pressure; LDL, low-density lipoprotein cholesterol; HDL, high-density lipoprotein cholesterol; TG, triglycerides; TC, total cholesterol; CRP, C-reactive protein.

Control function and stratification-based methods identify more overall causal effects than the TSLS methods, including all causal effects detected by linear TSLS. The quadratic control function approach identifies all but one relationship as nominally significant at the *p <* 0.05 level, including effects of all three exposures on age. All control function and stratification-based methods identify an overall effect of BMI on age, and PolyMR with model selection and residual stratification additionally identify an effect of alcohol consumption on age.

Among the TSLS-based methods, quadratic TSLS never rejects the linearity hypothesis, while NPIV with generalized B-spline polynomial basis identifies effects of BMI on all outcomes except for age as nonlinear (Table 2; Figure S7). None of the TSLS-based methods rejects the linearity hypothesis for age. In contrast, the control function and stratification-based methods identify many nonlinear signals, including in trait pairs where no overall effect is detected. In particular, quadratic control function identifies all but one effect as nonlinear, including effects of all three exposures on age. All control function and stratification-based methods identify nonlinear effects of BMI on age, and PolyMR and residual stratification further identify a nonlinear effect of alcohol consumption on age. The doubly-ranked stratification approach also reports a nonlinear effect of vitamin D on age despite finding no significant overall effect.

**Table 2:** Nonlinear Mendelian randomization results of nonlinearity inference for BMI, vitamin D, and alcohol consumption on blood pressure, lipid, C-reactive protein, and age. ★ indicates significant nonlinearity test results at *α* = 0.05, and —indicates non-significant results.

| Exposure | Outcome | TSLS-Q | NPIV-T | NPIV-B | CF-Q | PolyMR-D | PolyMR-S | Strat-RS |  | Strat-DR |  |
| --- | --- | --- | --- | --- | --- | --- | --- | --- | --- | --- | --- |
|  |  |  |  |  |  |  |  | Quad | Q | Quad | Q |
| BMI | SBP | — | — | ★ | ★ | ★ | ★ | ★ | ★ | ★ | ★ |
|  | DBP | — | — | ★ | ★ | ★ | ★ | ★ | ★ | ★ | ★ |
|  | LDL | — | — | ★ | ★ | ★ | ★ | ★ | ★ | ★ | ★ |
|  | HDL | — | ★ | ★ | ★ | ★ | ★ | ★ | ★ | ★ | ★ |
|  | TG | — | ★ | ★ | ★ | ★ | ★ | ★ | ★ | ★ | ★ |
|  | TC | — | ★ | ★ | ★ | ★ | ★ | ★ | ★ | — | ★ |
|  | CRP | — | ★ | ★ | ★ | ★ | ★ | ★ | ★ | ★ | ★ |
|  | Age | — | — | — | ★ | ★ | ★ | ★ | ★ | ★ | ★ |
| Vitamin D | SBP | — | — | — | ★ | — | ★ | — | — | — | — |
|  | DBP | — | — | — | ★ | — | — | — | — | — | — |
|  | LDL | — | — | — | ★ | ★ | — | ★ | ★ | — | — |
|  | HDL | — | — | — | ★ | ★ | ★ | ★ | — | ★ | — |
|  | TG | — | — | — | ★ | ★ | ★ | ★ | ★ | ★ | ★ |
|  | TC | — | — | — | ★ | ★ | — | ★ | ★ | — | — |
|  | CRP | — | — | — | — | — | ★ | — | ★ | — | — |
|  | Age | — | — | — | ★ | — | — | — | — | — | ★ |
| Alcohol | SBP | — | — | — | ★ | ★ | ★ | ★ | ★ | ★ | ★ |
|  | DBP | — | — | ★ | ★ | ★ | ★ | ★ | ★ | ★ | ★ |
|  | LDL | — | — | — | ★ | — | — | ★ | — | ★ | ★ |
|  | HDL | — | — | — | ★ | — | — | — | — | — | — |
|  | TG | — | — | — | ★ | — | — | ★ | — | ★ | — |
|  | TC | — | — | — | ★ | — | ★ | ★ | ★ | ★ | ★ |
|  | CRP | — | — | — | ★ | ★ | ★ | ★ | ★ | ★ | ★ |
|  | Age | — | — | — | ★ | — | ★ | — | ★ | — | — |

Abbreviations: TSLS-Q, quadratic two-stage least squares; NPIV-T, NPIV with tensor-product basis; NPIV-B, NPIV with generalized B-spline polynomial basis; CF-Q, quadratic control function; PolyMR-D, PolyMR without model selection (default); PolyMR-S, PolyMR with model selection; Strat-RS, residual stratification; Strat-DR, doubly-ranked stratification; Quad, quadratic test; Q, Cochran's Q test; SBP, systolic blood pressure; DBP, diastolic blood pressure; LDL, low-density lipoprotein cholesterol; HDL, high-density lipoprotein cholesterol; TG, triglycerides; TC, total cholesterol; CRP, C-reactive protein.

Among the TSLS-based methods, NPIV identifies an overall causal effect and rejects the linearity hypothesis for the effect of BMI on several traits. However, the fitted values from this method provide limited insights into the specific shapes of the causal relationships (Figures S8-S10). For example, for the effect of BMI on triglycerides (Figure S8), the NPIV point estimates suggest a positive effect at low BMI levels and a negative effect at moderate to high levels. However, the confidence bands cover zero across almost the entire range of BMI, making it difficult to draw definite conclusions. In contrast, quadratic control function suggests a similar pattern, but with substantially narrower confidence bands. The doubly-ranked stratification method also produces a fit with narrow confidence bands, suggesting that increases in BMI at moderate values raise triglyceride levels, whereas increases from already low or high BMI values have non-significant effects. However, based on simulation results, we do not expect these seemingly interpretable fits to be reliable given the overwhelming evidence for the presence of heterogeneity.

## 5 Discussion

In this study, we illustrate the impacts of heterogeneity of genetic associations with the exposure on estimates from nonlinear Mendelian randomization. Instrument-exposure association heterogeneity can induce substantial bias and inflate type I error in control function and stratification-based methods. In contrast, TSLS-based methods consistently control type I error but have low power to detect nonlinear effects or overall effects, especially under homogeneity. We further show that instrument-exposure effect heterogeneity can be identified using the Breusch-Pagan test. In UK Biobank analyses, we find widespread evidence of genetic effect heterogeneity for BMI, vitamin D, and alcohol consumption, potentially explaining previously reported spurious nonlinear findings involving these exposures[29, 48–50].

Our findings highlight an important gap in the current suite of nonlinear MR methods. Methods that are robust to heterogeneity are imprecise and underpowered, while methods that produce more precise estimates are vulnerable to false positives and bias in the presence of heterogeneity. This tradeoff is apparent in our UK Biobank analyses. In these settings, control function and stratification-based methods yield estimates with narrow confidence bands, but also produce false positives for the negative control outcome of age. This is consistent with our theoretical expectation that these methods are unreliable in the presence of genetic effect heterogeneity. By contrast, the more reliable TSLS methods avoid false positives, but also produce estimates with wide confidence bands, from which few scientific conclusions can be drawn.

This work has several limitations. First, our simulations do not fully capture realistic genome-wide architecture, as we only modeled independent causal variants and did not incorporate non-causal variants or linkage disequilibrium. However, we do not expect this to impact our primary conclusions. Second, we did not consider the effect of winner’s curse in instrument selection, as our design assumed independent discovery and evaluation cohorts. In practice, sample overlap, or the absence of external summary statistics may induce additional bias. Third, we focus on settings in which the sources of instrument-exposure effect heterogeneity are unobservable. Extending this framework to settings with observed effect modifiers and integrating this information through explicit interaction modeling may be valuable approaches.

## Data Availability

All data produced in the present study are available upon reasonable request to the authors.

## 7 Acknowledgments

This work was supported by National Human Genome Research Institute (NHGRI) R01 HG013104. This research has been conducted using the UK Biobank Resource under application number 24460.

## 8 Data and code availability

The code generated during this study are available at https://github.com/student-jw/nlmr_eval.

## Supplemental figures

**Figure S1:**
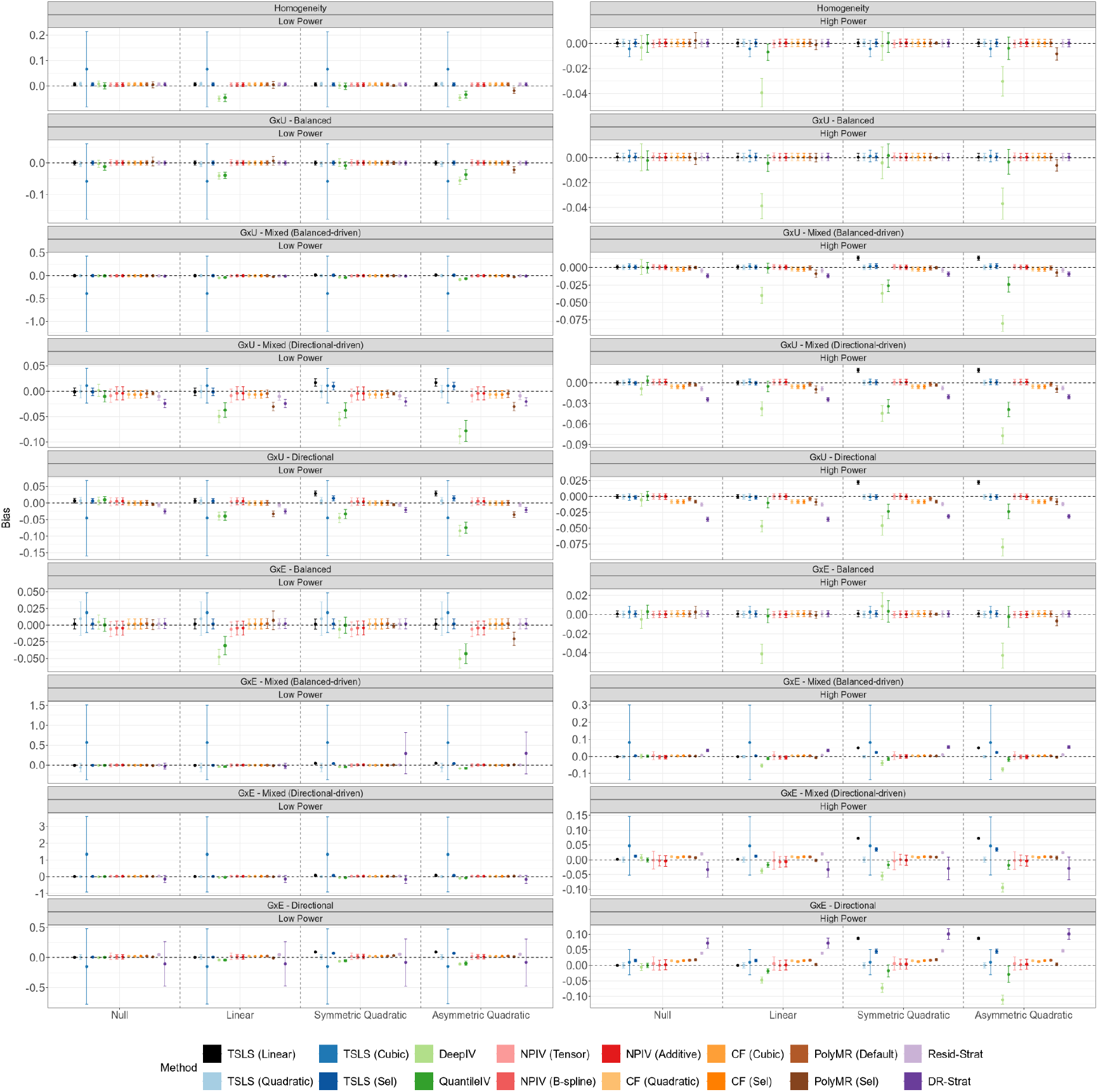
Average bias of nonlinear MR methods under instrument-exposure effect homogeneity or heterogeneity. Points indicate the average bias across 100 replicates, and error bars indicate 95% confidence intervals for the mean. Simulation sample size is 50, 000 (low power) or 100, 000 (high power). We varied the instrument-exposure effect structures across homogeneity, heterogeneity induced by *G* × *U*, and heterogeneity induced by *G* × *E*, each under balanced (*π* = 0), mixed (balanced-(*π* = 0.33) or directional-driven (*π* = 0.66)), and directional (*π* = 1) architectures. TSLS, two-stage least squares; NPIV, NPIV with tensor-product basis (Tensor), generalized B-spline polynomial basis (B-spline), or additive basis (Additive); CF, control function; PolyMR (Default), PolyMR without model selection; PolyMR (Sel), PolyMR with model selection; Resid-Strat, residual stratification; DR-Strat, doubly-ranked stratification.

**Figure S2:**
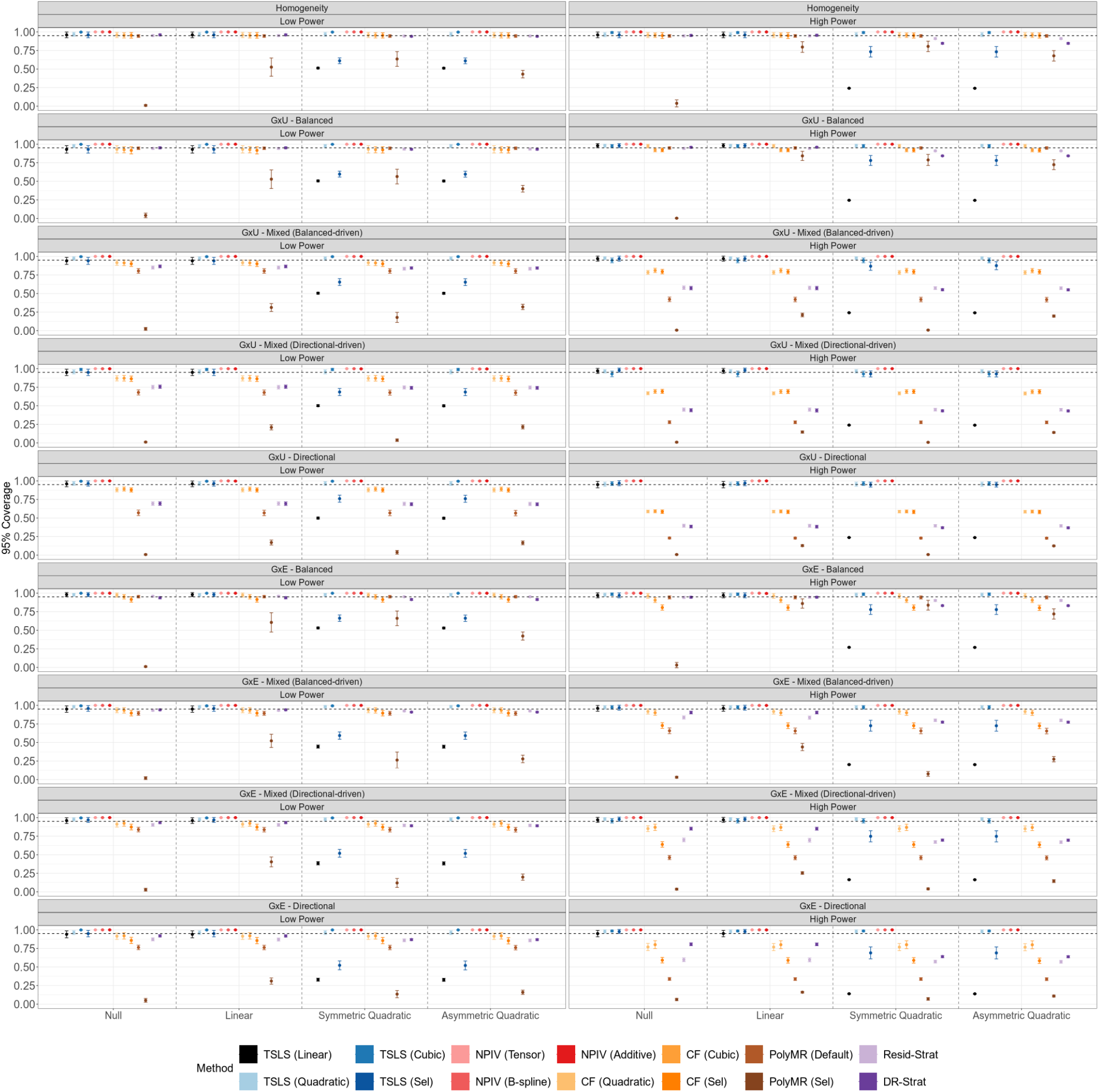
Average 95% confidence interval coverage of nonlinear MR methods under instrument-exposure effect homogeneity or heterogeneity. Points indicate the average coverage across 100 replicates, and error bars indicate 95% confidence intervals for the mean. Simulation sample size is 50, 000 (low power) or 100, 000 (high power). We varied the instrument-exposure effect structures across homogeneity, heterogeneity induced by *G* × *U*, and heterogeneity induced by *G* × *E*, each under balanced (*π* = 0), mixed (balanced-(*π* = 0.33) or directional-driven (*π* = 0.66)), and directional (*π* = 1) architectures. TSLS, two-stage least squares; NPIV, NPIV with tensor-product basis (Tensor), generalized B-spline polynomial basis (B-spline), or additive basis (Additive); CF, control function; PolyMR (Default), PolyMR without model selection; PolyMR (Sel), PolyMR with model selection; Resid-Strat, residual stratification; DR-Strat, doubly-ranked stratification.

**Figure S3:**
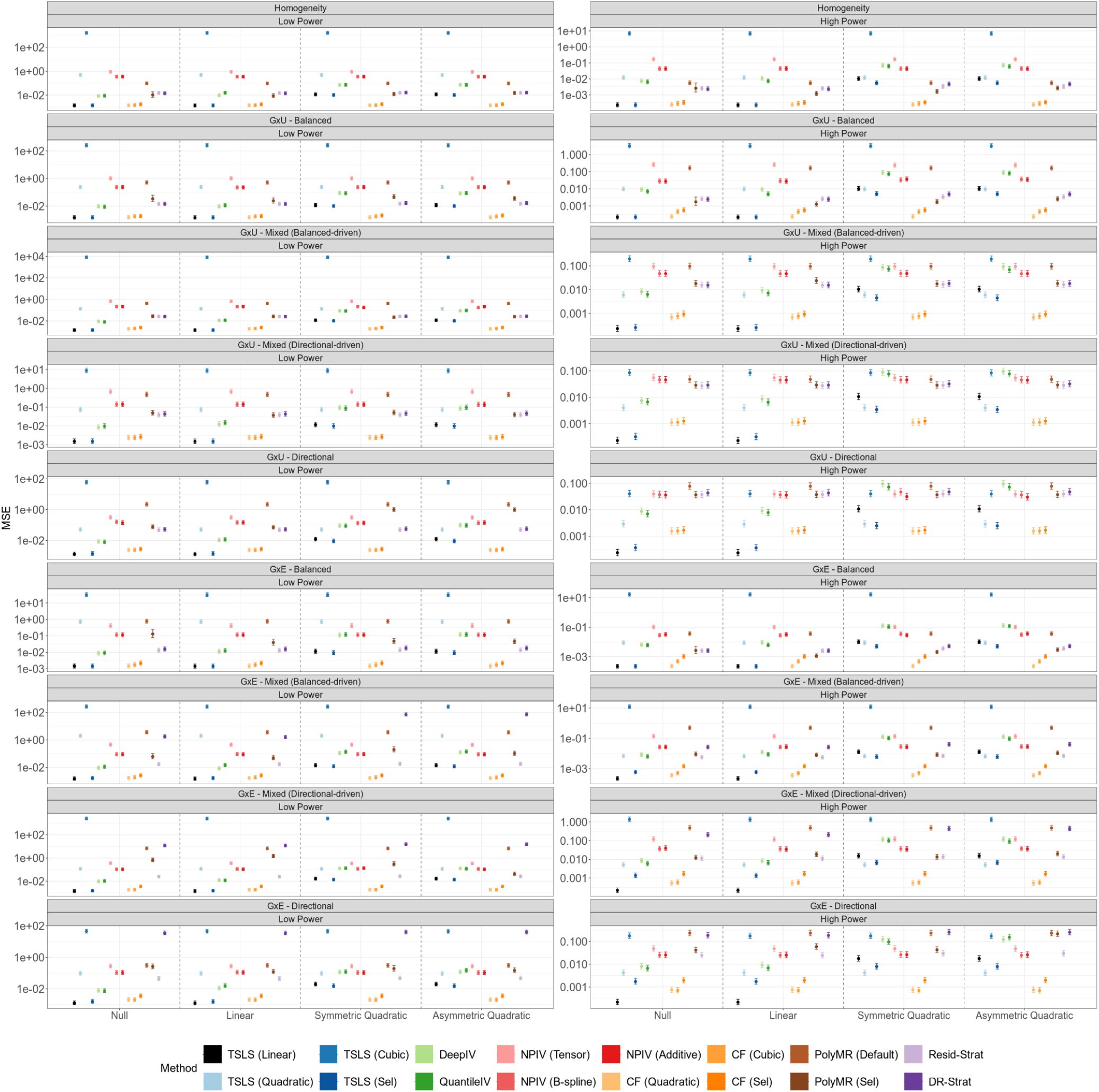
Average mean squared error (MSE) of nonlinear MR methods under instrument-exposure effect homogeneity or heterogeneity. Points indicate the average MSE across 100 replicates, and error bars indicate 95% confidence intervals for the mean. Simulation sample size is 50, 000 (low power) or 100, 000 (high power). We varied the instrument-exposure effect structures across homogeneity, heterogeneity induced by *G* × *U*, and heterogeneity induced by *G* × *E*, each under balanced (*π* = 0), mixed (balanced-(*π* = 0.33) or directional-driven (*π* = 0.66)), and directional (*π* = 1) architectures. TSLS, two-stage least squares; NPIV, NPIV with tensor-product basis (Tensor), generalized B-spline polynomial basis (B-spline), or additive basis (Additive); CF, control function; PolyMR (Default), PolyMR without model selection; PolyMR (Sel), PolyMR with model selection; Resid-Strat, residual stratification; DR-Strat, doubly-ranked stratification.

**Figure S4:**
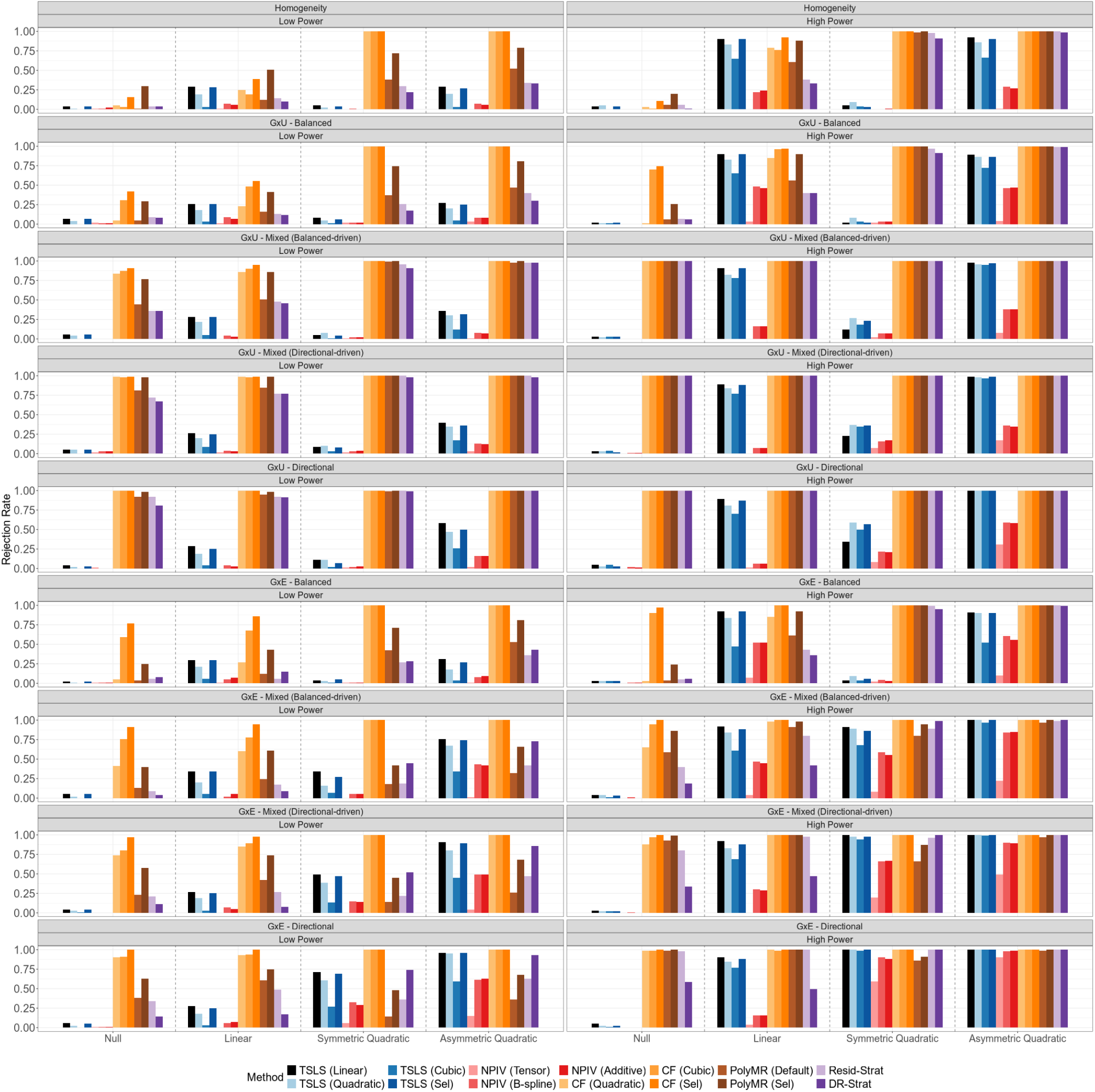
Rejection rates for overall-effect testing of nonlinear MR methods. Rejection rates were calculated from *N* = 50, 000 or 100, 000 simulated individuals (low-versus high-power settings) per replicate, and summarized across 100 replicates. We varied the instrument-exposure effect structures across homogeneity, heterogeneity induced by *G* × *U*, and heterogeneity induced by *G* × *E*, each under balanced (*π* = 0), mixed (balanced-(*π* = 0.33) or directional-driven (*π* = 0.66)), and directional (*π* = 1) architectures. TSLS, two-stage least squares; NPIV, NPIV with tensor-product basis (Tensor), generalized B-spline polynomial basis (B-spline), or additive basis (Additive); CF, control function; PolyMR (Default), PolyMR without model selection; PolyMR (Sel), PolyMR with model selection; Resid-Strat, residual stratification; DR-Strat, doubly-ranked stratification.

**Figure S5:**
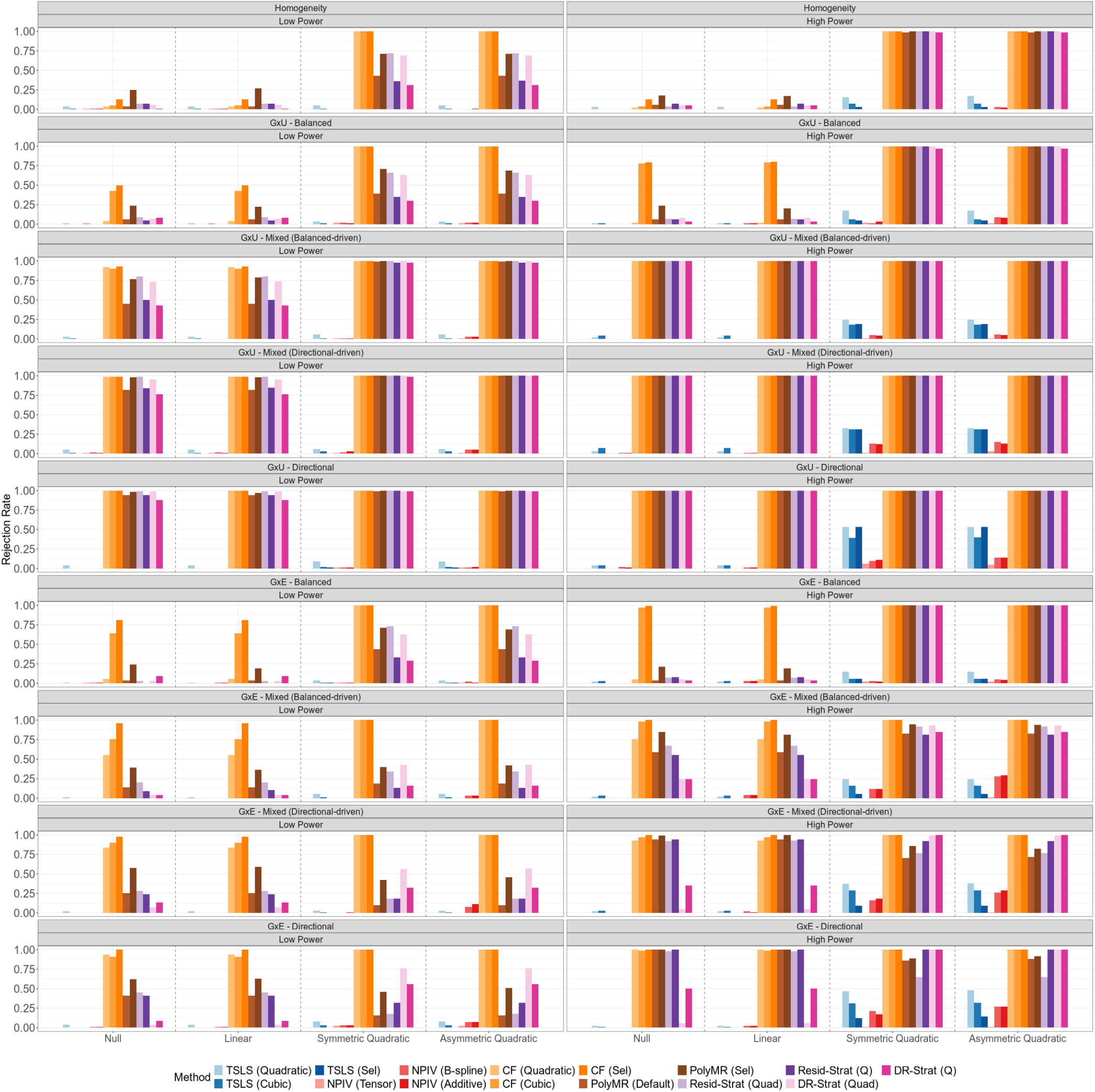
Rejection rates for nonlinearity testing of nonlinear MR methods. Rejection rates were calculated from *N* = 50, 000 or 100, 000 simulated individuals (low-versus high-power settings) per replicate, and summarized across 100 replicates. We varied the instrument-exposure effect structures across homogeneity, heterogeneity induced by *G* × *U*, and heterogeneity induced by *G* × *E*, each under balanced (*π* = 0), mixed (balanced-(*π* = 0.33) or directional-driven (*π* = 0.66)), and directional (*π* = 1) architectures. TSLS, two-stage least squares; NPIV, NPIV with tensor-product basis (Tensor), generalized B-spline polynomial basis (B-spline), or additive basis (Additive); CF, control function; PolyMR (Default), PolyMR without model selection; PolyMR (Sel), PolyMR with model selection; Resid-Strat, residual stratification; DR-Strat, doubly-ranked stratification; Quad, quadratic test; Q, Cochran’s Q test.

**Figure S6:**
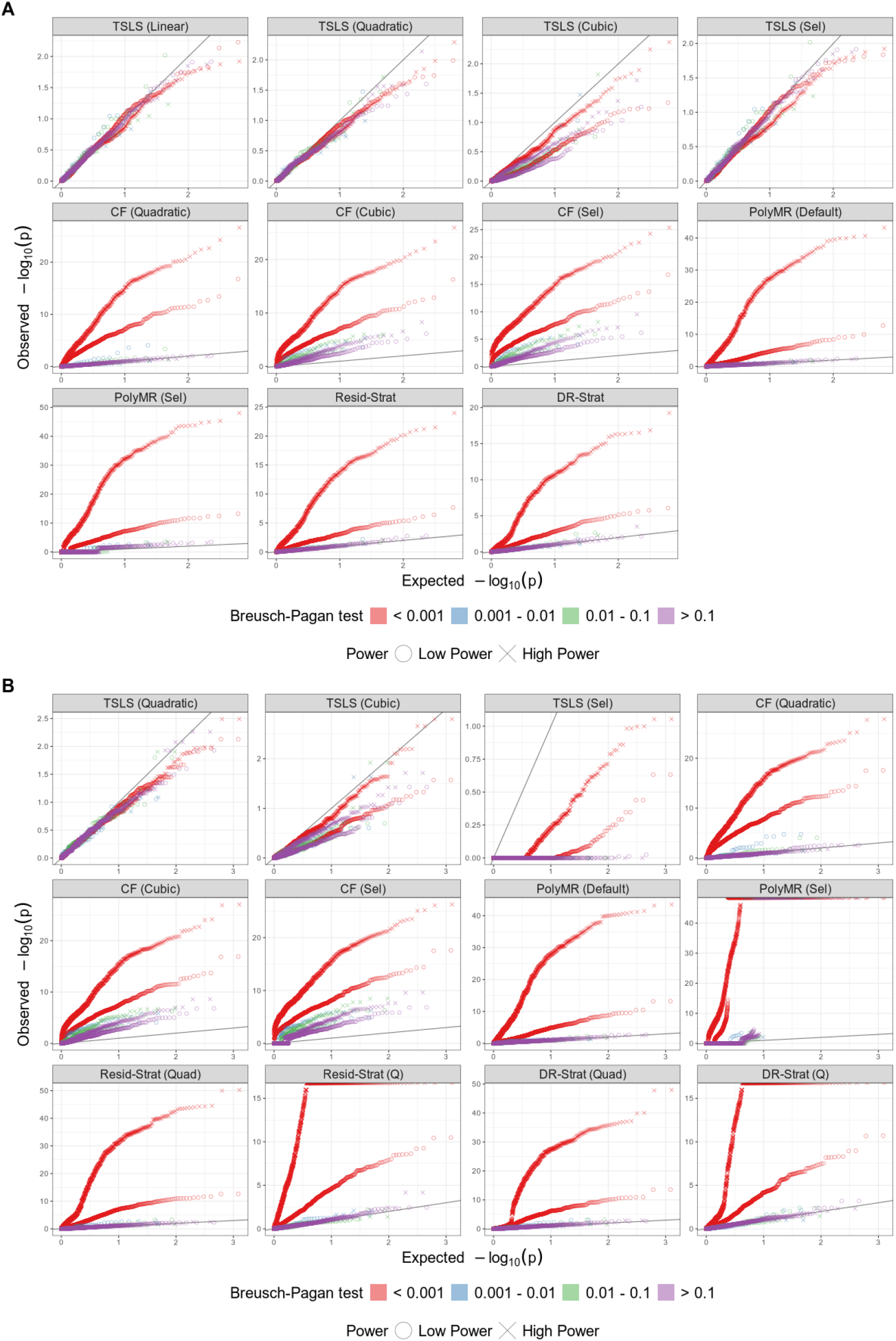
Quantile–Quantile plots for overall-effect and nonlinearity testing of nonlinear MR methods. (A) Overall-effect testing, evaluated under the null hypothesis of no causal effect. (B) Nonlinearity testing, evaluated under the null hypothesis of no causal effect or a linear effect. P-values were evaluated from *N* = 50, 000 or 100, 000 simulated individuals (low-versus high-power settings), and pooled across all homogeneity and heterogeneity scenarios over 100 replicates. TSLS, two-stage least squares; CF, control function; PolyMR (Default), PolyMR without model selection; PolyMR (Sel), PolyMR with model selection; Resid-Strat, residual stratification; DR-Strat, doubly-ranked stratification; Quad, quadratic test; Q, Cochran’s Q test.

**Figure S7:**
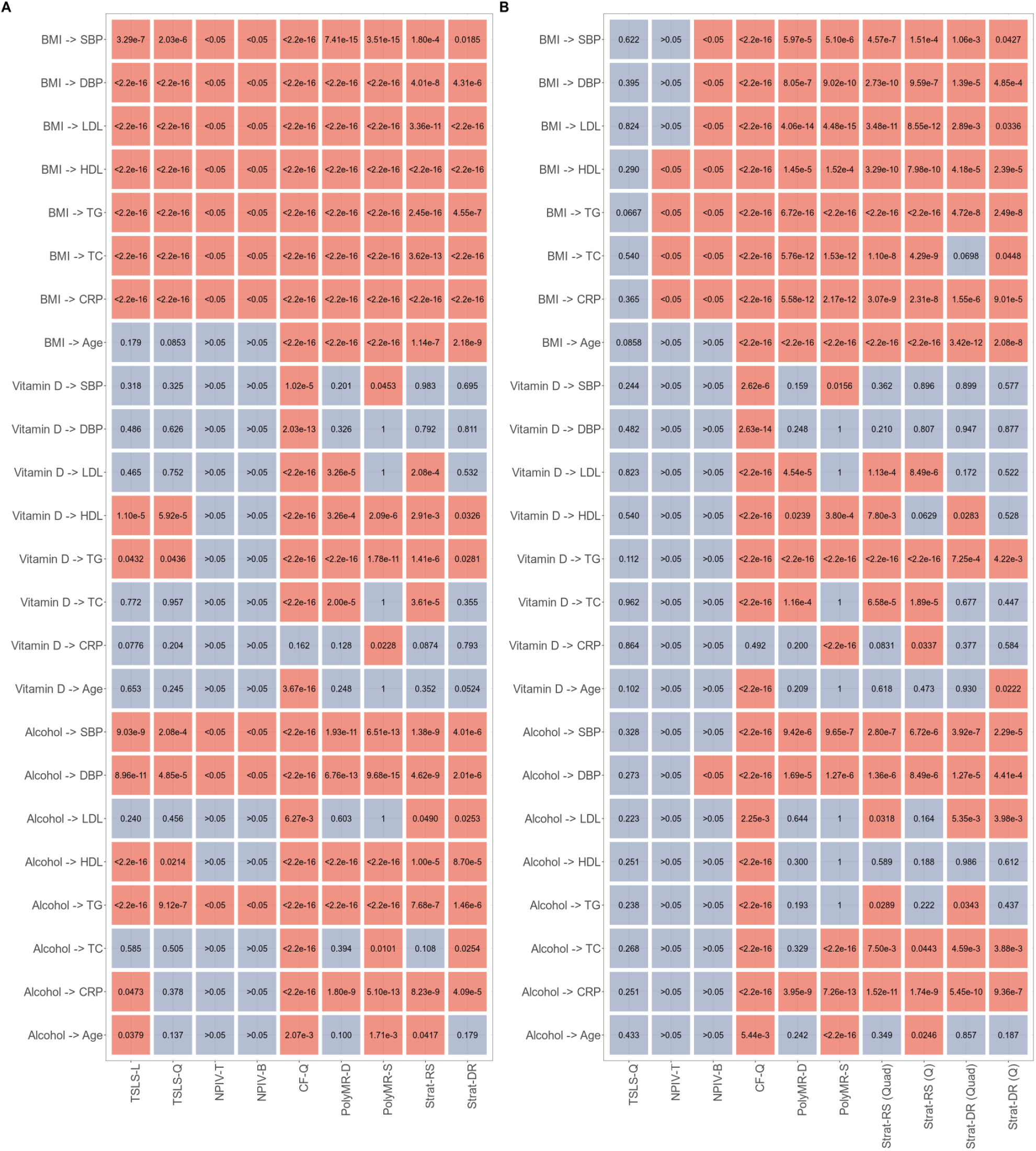
Mendelian randomization results for overall-effect and nonlinearity inference of BMI, vitamin D, and alcohol consumption on blood pressure, lipid, C-reactive protein, and age in UK Biobank. (A) Overall-effect testing. (B) Nonlinearity testing. Numbers denote p-values for the corresponding test; red indicates nominal significance at *α* = 0.05 and blue indicates non-significant results. TSLS-L, linear two-stage least squares; TSLS-Q, quadratic two-stage least squares; NPIV-T, NPIV with tensor-product basis; NPIV-B, NPIV with generalized B-spline polynomial basis; CF-Q, quadratic control function; PolyMR-D, PolyMR without model selection (default); PolyMR-S, PolyMR with model selection; Strat-RS, residual stratification; Strat-DR, doubly-ranked stratification; Quad, quadratic test; Q, Cochran’s Q test; SBP, systolic blood pressure; DBP, diastolic blood pressure; LDL, low-density lipoprotein cholesterol; HDL, high-density lipoprotein cholesterol; TG, triglycerides; TC, total cholesterol; CRP, C-reactive protein.

**Figure S8:**
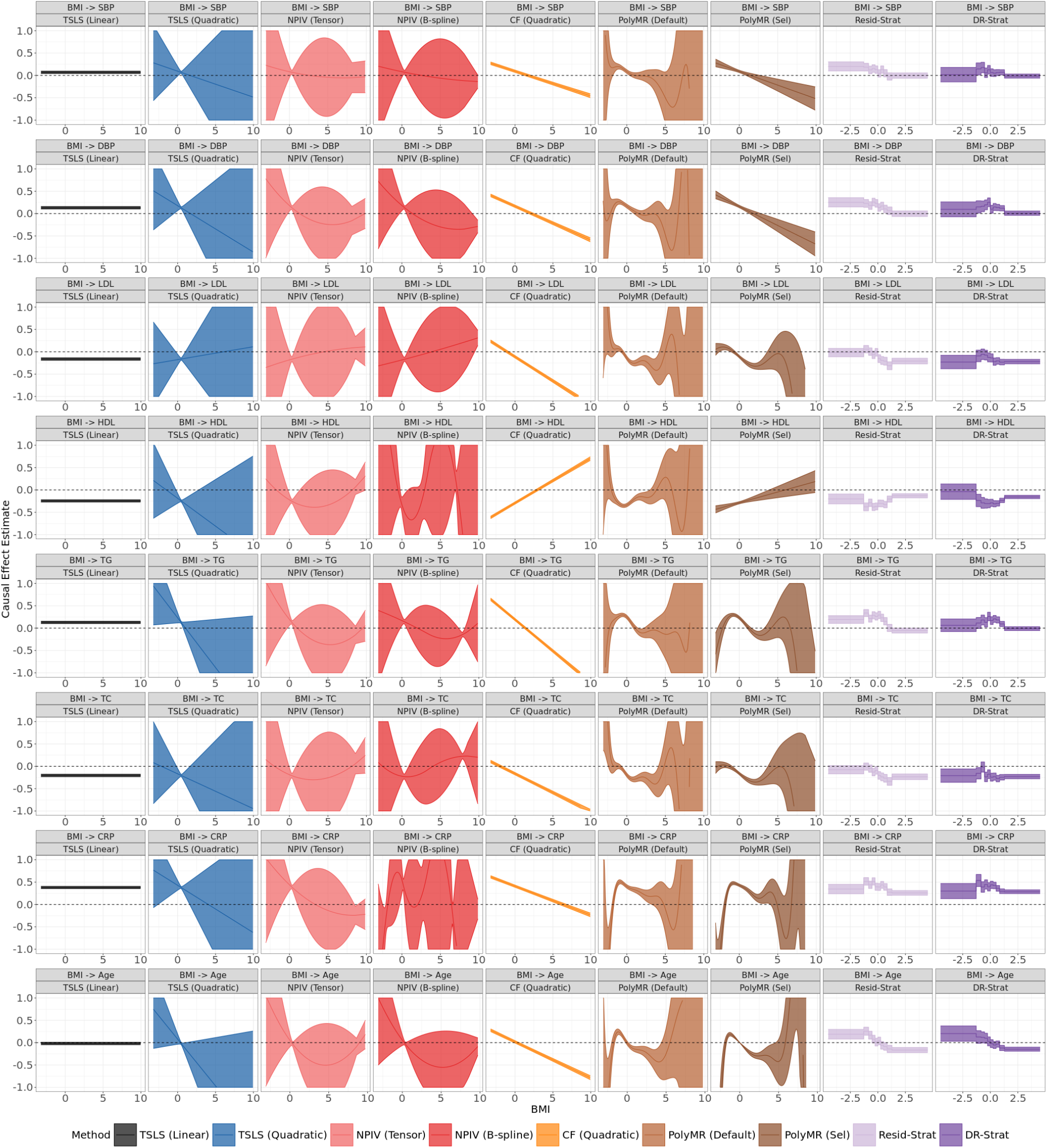
Estimated causal shapes of BMI on blood pressure, lipid, C-reactive protein, and age in UK Biobank. Shaded regions indicate 95% confidence bands. The x-axis denotes standardized BMI, except for stratification-based methods, where it represents rank-based inverse-normal-transformed post-stratification BMI. TSLS, two-stage least squares; NPIV, NPIV with tensor-product basis (Tensor), generalized B-spline polynomial basis (B-spline); CF, control function; PolyMR (Default), PolyMR without model selection; PolyMR (Sel), PolyMR with model selection; Resid-Strat, residual stratification; DR-Strat, doubly-ranked stratification; SBP, systolic blood pressure; DBP, diastolic blood pressure; LDL, low-density lipoprotein cholesterol; HDL, high-density lipoprotein cholesterol; TG, triglycerides; TC, total cholesterol; CRP, C-reactive protein.

**Figure S9:**
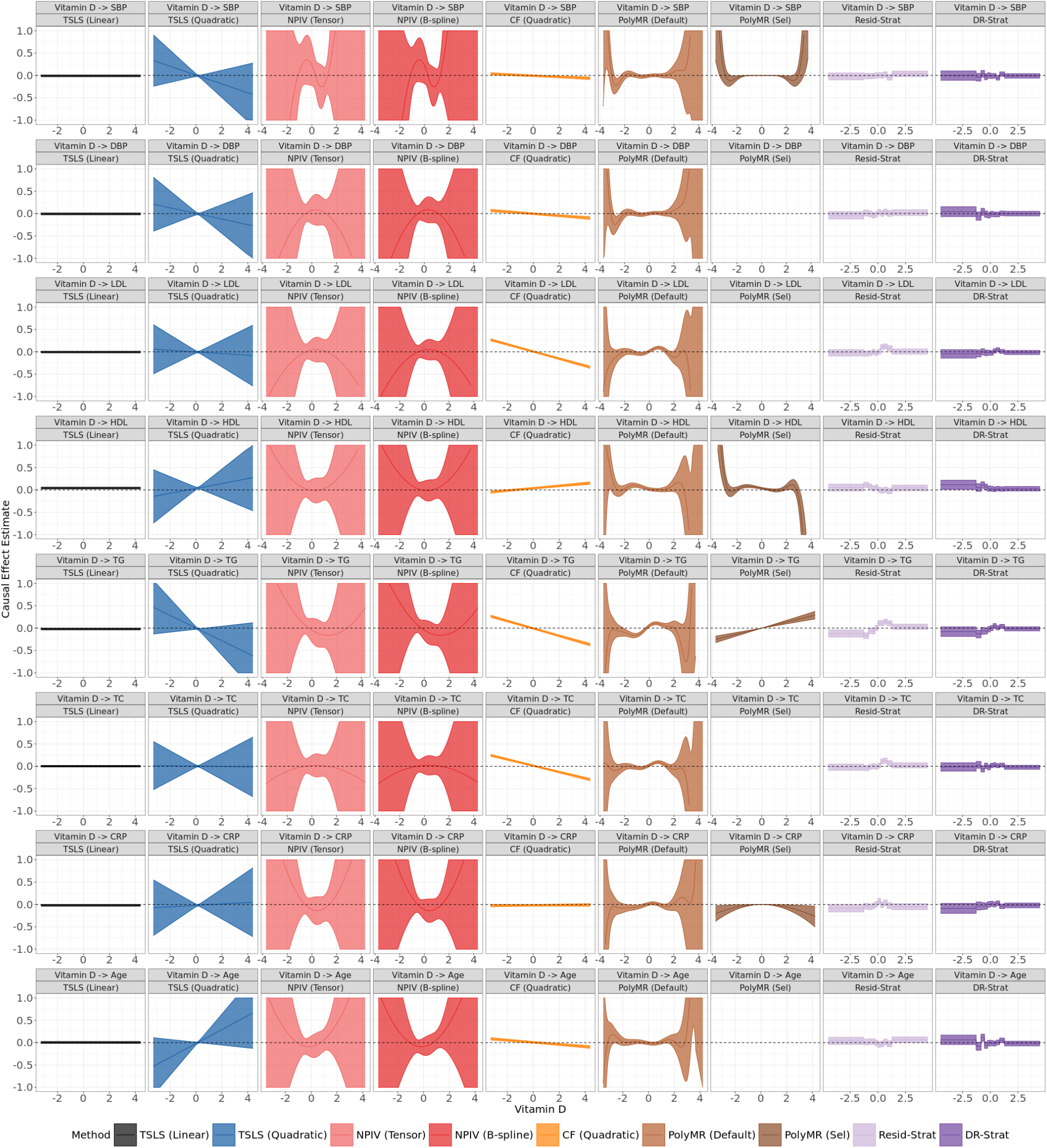
Estimated causal shapes of vitamin D on blood pressure, lipid, C-reactive protein, and age in UK Biobank. Shaded regions indicate 95% confidence bands. The x-axis denotes standardized vitamin D (natural-log scale), except for stratification-based methods, where it represents rank-based inverse-normal-transformed post-stratification vitamin D (natural-log scale). TSLS, two-stage least squares; NPIV, NPIV with tensor-product basis (Tensor), generalized B-spline polynomial basis (B-spline); CF, control function; PolyMR (Default), PolyMR without model selection; PolyMR (Sel), PolyMR with model selection; Resid-Strat, residual stratification; DR-Strat, doubly-ranked stratification; SBP, systolic blood pressure; DBP, diastolic blood pressure; LDL, low-density lipoprotein cholesterol; HDL, high-density lipoprotein cholesterol; TG, triglycerides; TC, total cholesterol; CRP, C-reactive protein.

**Figure S10:**
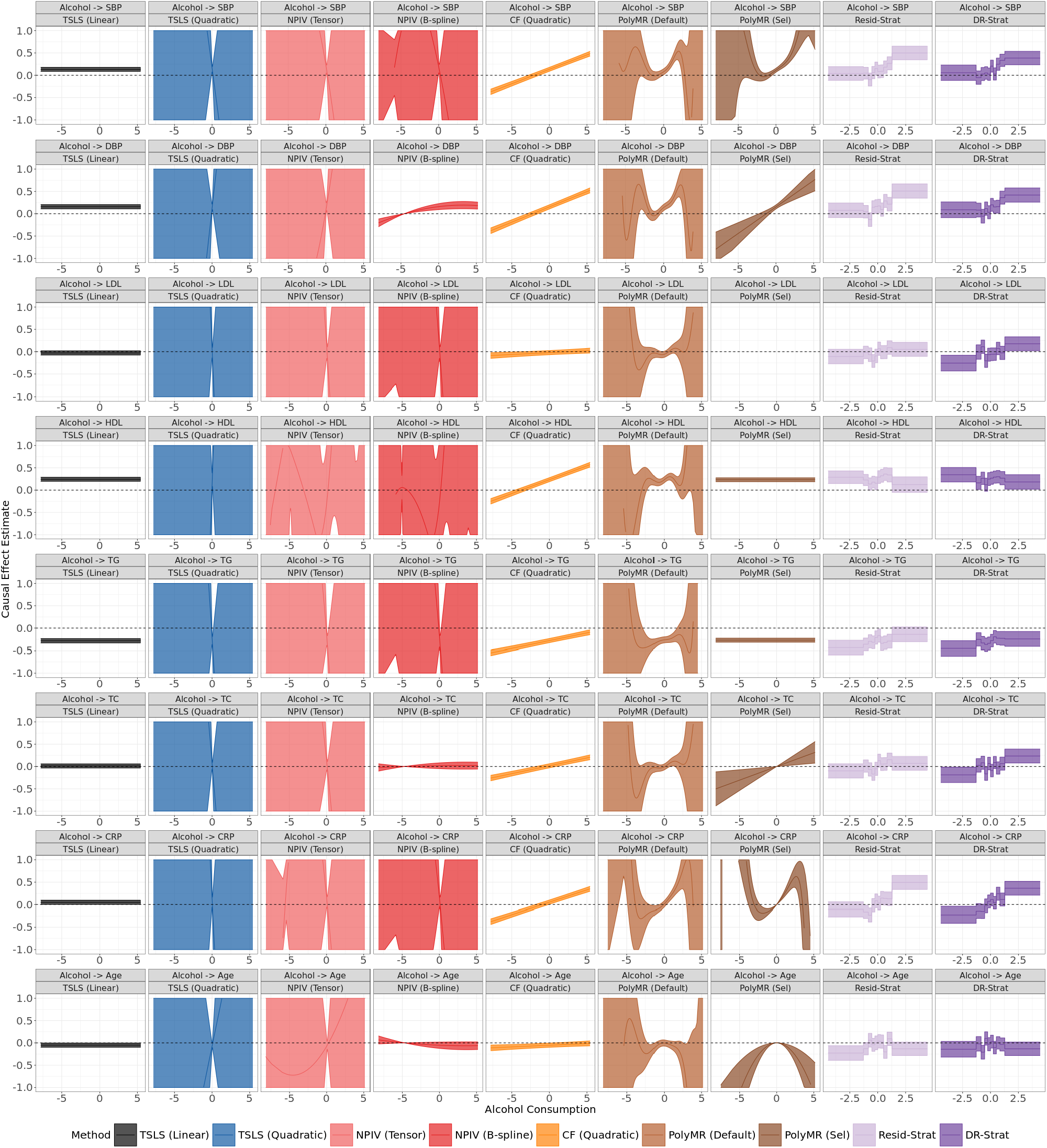
Estimated causal shapes of alcohol consumption on blood pressure, lipid, C-reactive protein, and age in UK Biobank. Shaded regions indicate 95% confidence bands. The x-axis denotes standardized alcohol consumption (natural-log scale), except for stratification-based methods, where it represents rank-based inverse-normal-transformed post-stratification alcohol consumption (natural-log scale). TSLS, two-stage least squares; NPIV, NPIV with tensor-product basis (Tensor), generalized B-spline polynomial basis (B-spline); CF, control function; PolyMR (Default), PolyMR without model selection; PolyMR (Sel), PolyMR with model selection; Resid-Strat, residual stratification; DR-Strat, doubly-ranked stratification; SBP, systolic blood pressure; DBP, diastolic blood pressure; LDL, low-density lipoprotein cholesterol; HDL, high-density lipoprotein cholesterol; TG, triglycerides; TC, total cholesterol; CRP, C-reactive protein.

## Supplemental notes

### Note S1: Details of nonlinear MR method implementation

We implemented Deep IV and Quantile IV using the `ml-mr` Python package, with the `--exposure-network-type gaussian_net` option for Deep IV and the `--n-quantiles 10` option for Quantile IV. NPIV was implemented using the `npiv` R package, with three basis specifications: (1) tensor-product basis (default), (2) generalized B-spline polynomial basis (`basis = “glp”`), and (3) additive basis (`basis = “additive”`). PolyMR was implemented using the `PolyMR` R package, both without model selection (default) and with model selection (`p_thr_drop = NULL`). Residual stratification and doubly-ranked stratification were implemented using the `SUMnlmr` R package.

